# CLEAR: An Auditable Foundation Model for Radiology Grounded in Clinical Concepts

**DOI:** 10.64898/2026.01.15.26344222

**Authors:** Tianyu Han, Riga Wu, Yu Tian, Firas Khader, Lisa C. Adams, Keno K. Bressem, Christos Davatzikos, Jakob Nikolas Kather, Li Shen, David A. Mankoff, Eduardo Mortani Barbosa, Daniel Truhn

## Abstract

“Black box” deep learning models for medical image interpretation limit clinical trust and analysis of performance degradation. Here, we introduce Concept-Level Embeddings for Auditable Radiology (CLEAR), an auditable foundation model based on clinical concepts. Trained on over 0.87 million image-report pairs from 239,091 patients, CLEAR learns a visual representation and projects chest X-rays into a semantically rich space defined by large language model embeddings, making every prediction traceable to specific radiological observations. External validation on four large, physician-annotated datasets from the United States, Europe, and Asia shows that CLEAR not only achieves state-of-the-art classification performance but also enables novel applications: auditable zero-shot pathology detection, systematic identification of radiological confounders, and the creation of expert-level concept bottleneck models from data-driven concepts. By integrating clinical knowledge directly into its reasoning process, CLEAR offers a framework for robust model auditing, safer deployment, and enhanced physician-AI collaboration, advancing towards trustworthy medical AI.

## Main

Medical imaging errors cause an estimated 40,000 to 80,000 preventable deaths annually in the U.S., with radiological misdiagnosis affecting up to 1 in 10 patients.^1, 2^ Diagnostic imaging accounts for over 100 billion dollar in annual spending, yet interpretation errors drive repeat scans, delayed treatment, and malpractice costs exceeding 4 billion dollar each year.^3^ Significant progress in computer-assisted diagnosis, particularly methods leveraging end-to-end machine learning (ML), has been made across radiological tasks, including disease progression prediction,^4, 5, 6, 7, 8, 9, 10^ tissue segmentation,^11, 12, 13, 14, 15^ findings classification,^16, 17, 18, 19, 20, 21^ survival prediction,^22, 23^ medical report generation,^24, 25, 26, 27^ and multimodal situation awareness.^28, 29, 30, 31, 32, 33, 34, 35^ In addition, foundation models can use large cohorts of unlabeled training images to address a wide range of clinical tasks, such as diagnosing rare (long-tailed) thoracic diseases^21^ and predicting cancer imaging biomarkers.^36^

However, the decision-making processes and predictive features of these models remain largely uninterpretable to even experienced radiologists. More importantly, they are not necessarily grounded in established clinical concepts or biological processes.^37, 38, 39, 40, 41^ This lack of transparency is not merely an academic concern. As a result, they are vulnerable to learning spurious correlations from confounders in the data, making training via computational approaches like gradient back-propagation inherently unreliable.^39, 42, 43, 44, 45, 46^ Consequently, the generalization of even state-of-the-art foundation models must often be taken on faith, with little to no insight into the underlying causes of performance degradation. This stands in stark contrast to the practice of radiologists, who rely on written language to interpret and communicate radiological findings in reports to patients and clinical teams.

To facilitate auditing within the target application, inherently interpretable models like concept bottleneck models (CBMs),^47^ LaBo,^48^ CEM,^49^ and post-hoc CBM,^50^ use domain-specific, expert-annotated concepts to guide predictions. In biomedicine, CBMs have been applied to tasks such as skin lesion classification,^39, 51^ choroid neoplasias diagnosis,^52^ neurological disorder diagnosis,^53^ and medical image retrieval.^54^ However, CBMs have important limitations: the choice of concepts often reflects the subjectivity and constraints of individual reporters and thus may be neither comprehensive nor optimal for the predictive task. Moreover, the sets are not easily scalable and require dense concept annotations in the training set. CBMs generally demonstrate lower performance than end-to-end models and have not been successfully applied to zero-shot image classification in medicine.^39, 47^ Addressing these limitations is fundamental to building trustworthy models for medical image interpretation that are both generalizable and applicable to routine clinical practice on a broad scale.

In this study, we introduce Concept-Level Embeddings for Auditable Radiology (CLEAR), which leverages the collective knowledge of the radiological community, as encapsulated in publicly available radiological reports (Figure 1a), to construct a comprehensive and semantically rich concept space for CXR interpretation using 368,294 large language model (LLM)-embedded radiological observations. CLEAR was developed using over 0.87 million image-report pairs from the publicly available MIMIC-CXR,^55^ CheXpert-Plus,^56^ and ReXGradient^57^ datasets through task-agnostic contrastive pretraining (Figure 1b). We first trained a DINOv2 image encoder^58^ and a corresponding text encoder^59^ to map visual and textual information into a shared semantic space. At inference, these encoders work in tandem: the model assigns a score to each input CXR for a given radiological observation by measuring the similarity between the outputs of the image encoder and the text encoder, thereby indicating the degree to which the image represents that observation (Figure 1c). The resulting vector of similarity scores is then transferred into the semantically rich embedding space of a state-of-the-art LLM, such as OpenAI’s text-embedding-3-small (2025), Qwen3-embedding-8B (2025),^60, 61^ SFR-embedding-Mistral (2024),^62^ or BiomedBERT (2021)^63^ (Figure 1d and Figure S1c), through matrix multiplication, where the vector of similarity scores is multiplied by the pre-computed LLM embeddings for all radiological observations (Figure 1d). This final, concept-based embedding takes advantage of the LLM’s ability to capture nuanced semantic relationships within complex medical text, allowing precise interpretation and comparison. We demonstrate CLEAR’s superior zero-shot performance and its capability for auditing zero-shot classification across a wide array of tasks, including classification of cardiac, pulmonary, bone, and vascular findings, using four external CXR benchmarks from the United States, Spain, and Vietnam. CLEAR outperforms other vision-language foundation models, including CheXzero,^20^ BiomedCLIP,^64^ and OpenAICLIP,^65^ on task-specific supervised probing. We also demonstrate that both the standard CBM and our CLEAR model achieve expert-level pathology detection on the CheXpert competition using high-quality concepts discovered by our data-driven approach, without sacrificing interpretability.

**Figure 1:**
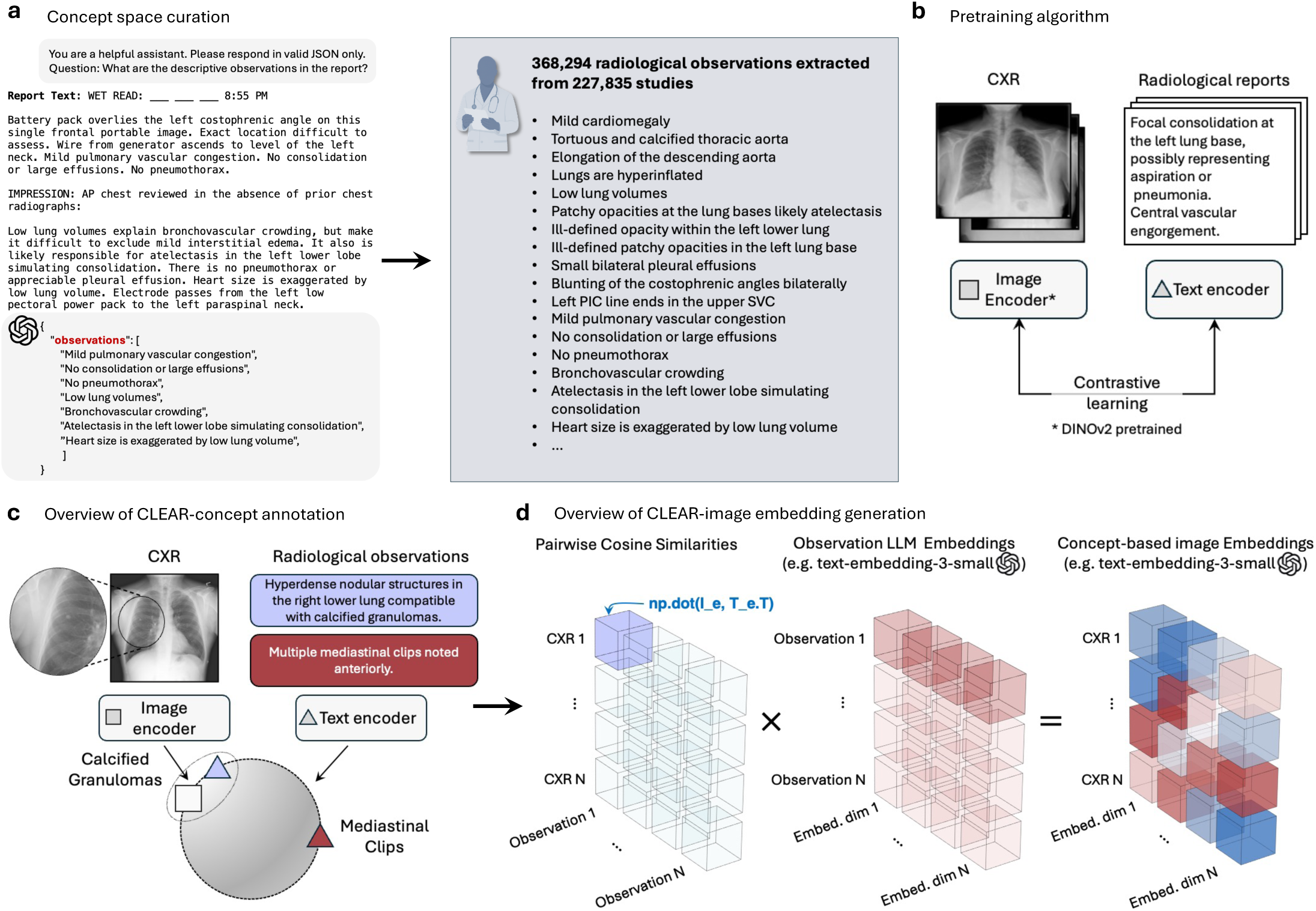
The CLEAR framework. **(a)** Concept space curation: An LLM extracted 368,294 radiological observations from the report text of 227,835 studies, including findings such as “mild cardiomegaly”, “tortuous and calcified thoracic aorta”, “elongation of the descending aorta”, and various other clinical observations ranging from lung pathologies to cardiac findings. **(b)** Pretraining algorithm: CXR and radiological reports are processed through an image encoder (DINOv2 pretrained) and text encoder connected via contrastive learning to align visual and textual representations. **(c)** Overview of CLEAR-concept annotation: Radiological observations are mapped to specific image features. **(d)** Overview of CLEAR-image embedding generation: CXRs and observations are projected into a shared semantic space through three stages: pairwise cosine similarities, LLM-generated observation embeddings (e.g., SFR-Embedding-Mistral), and final concept-based image embeddings. This unified representation enables direct comparison between radiological images and clinical concepts through matrix multiplication of similarity scores and observation embeddings.

## Results

### Auditable zero-shot classification of diverse findings

CLEAR performs zero-shot classification by first encoding each CXR as a vector of similarity scores between the image and 368,294 clinical observations (concepts) extracted from radiology reports, computed via pairwise dot products. This image-concept score vector quantifies the extent to which the CXR expresses each clinical concept and is subsequently projected into an LLM embedding space (such as Salesforce’s SFR-Embedding-Mistral) by multiplying it by the corresponding concept embeddings, resulting in a concept-based image embedding. For zero-shot classification, CLEAR constructs a pair of prompts for each disease or clinical finding: a positive prompt (e.g., “atelectasis”) and a negative prompt (e.g., “no atelectasis”). Encoding both prompts using the same LLM embedding model^66^ ensures that both the image embedding and prompt embeddings reside in a unified semantic space. The model then computes logits by measuring similarities between the image embedding and each prompt pair. Finally, a softmax function applied to these paired logits produces a probability score that directly indicates the likelihood of the specific disease or finding being present, enabling multi-label classification where multiple pathologies may coexist (Figure 2a).

**Figure 2:**
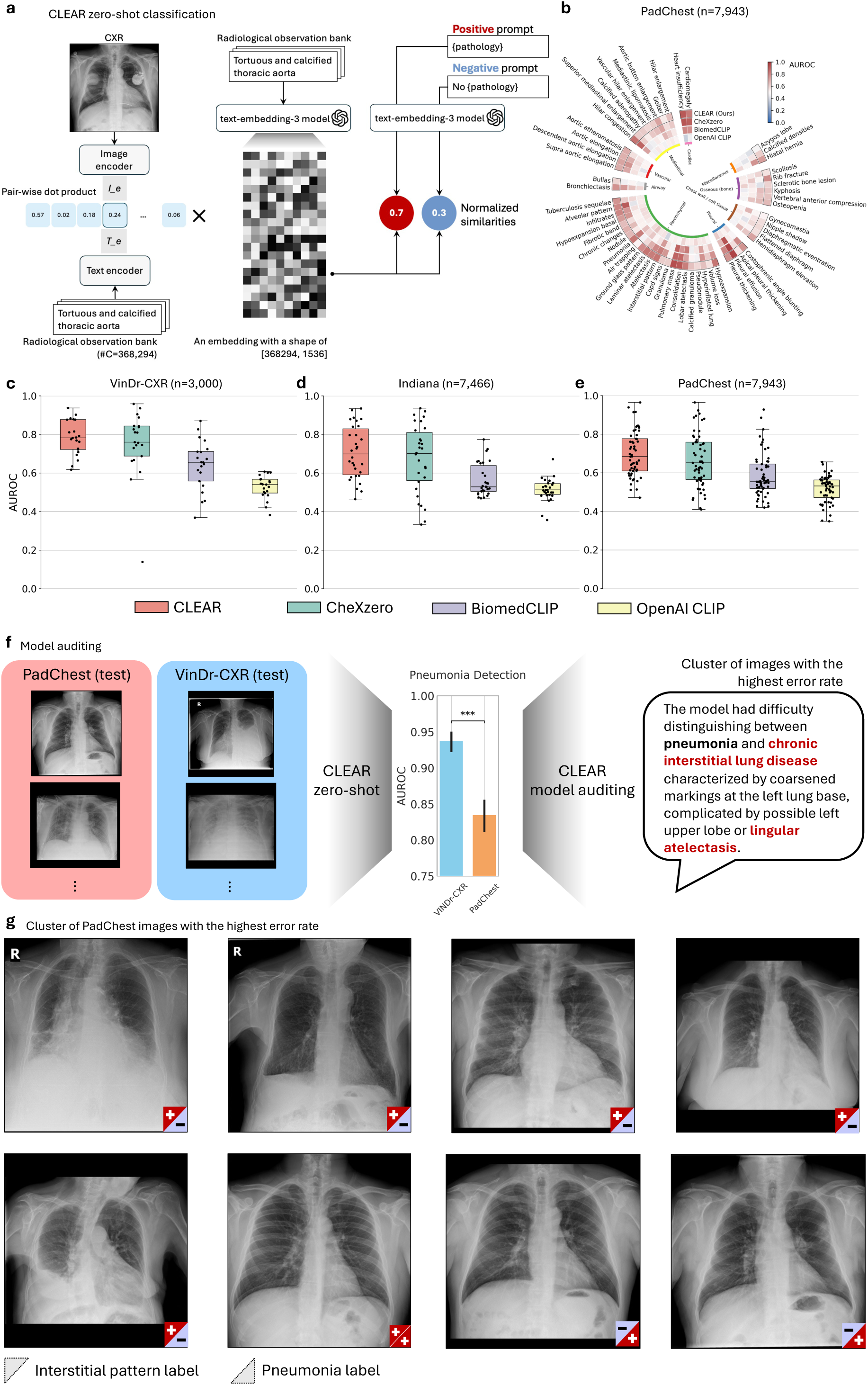
Zero-shot classification and auditing capabilities of CLEAR. **(a)** Schematic of the CLEAR zero-shot classification pipeline. CXRs are processed through an image encoder to compute pairwise dot products with text-encoded radiological observations from a bank of 368,294 concepts. The resulting similarity scores are multiplied by LLM-encoded observation embeddings (e.g., text-embedding-3 model or SFR-Embedding-Mistral model) to create concept-based image embeddings. Classification is performed by computing normalized similarities between the image embedding and positive/negative prompt pairs for each pathology. **(b)** Circular heatmap showing zero-shot AUROC performance on the PadChest dataset across 55 phenotype findings. Color intensity represents AUROC values from 0 (blue) to 1.0 (red), with CLEAR outperforming comparison models. **(c-e)** Zero-shot performance comparison across external datasets. Box plots show AUROC distributions for CLEAR, CheXzero, BiomedCLIP, and OpenAICLIP on **(c)**, VinDr-CXR (n=3,000; 21 findings), **(d)**, Indiana (n=7,466; 30 findings), and **(e)**, PadChest (n=7,943; 55 findings). Boxes represent interquartile ranges with median lines; whiskers extend to 1.5× interquartile range. **(f)** Model auditing reveals performance disparities in pneumonia detection between the VinDr-CXR (AUROC=0.93) and PadChest (AUROC=0.83) test sets. CLEAR identifies that the highest error rate cluster contains images with interstitial lung disease and lingular atelectasis, known confounders that can mask pneumonia-related consolidation. **(g)** Representative CXRs from the PadChest cluster with the highest pneumonia detection error rate. The true interstitial pattern and true pneumonia labels for each image are represented by the color and symbols in the upper left and lower right triangles in the small box, respectively.

We externally evaluated CLEAR on three physician-annotated CXR datasets: VinDr-CXR (n=3,000; 21 findings), PadChest (n=7,943; 55 findings), and the Indiana University CXR Collection (Indiana; n=7,466; 30 findings). For all evaluations, the area under the receiver operating characteristic curve (AUROC) was used as the primary performance metric to compare CLEAR with state-of-the-art vision-language foundation models, including CheXzero, BiomedCLIP, and OpenAICLIP. Detailed performance results are provided in Supplementary Tables S1–S4, and dataset descriptions can be found in the Methods section.

On the VinDr-CXR benchmark (Figure 2c), CLEAR achieved an average zero-shot AUROC of 78.2%, outperforming the next-best model, CheXzero (mean AUROC 75.0%), with statistically significant improvements (*P <* 0.01, two-sided paired permutation test) on findings such as “nodule/mass”, “lung tumor”, “pulmonary fibrosis”, and “no finding”. On the Indiana benchmark (Figure 2d), CLEAR outperformed CheXzero on 10 findings and underperformed on 2 (*P <* 0.01); outperformed BiomedCLIP on all 22 findings); and outperformed OpenAICLIP on all 20 findings. On the PadChest benchmark (Figure 2b and e), CLEAR achieved a mean AUROC of 70.0%, outperforming CheXzero (66.8%), BiomedCLIP (59.2%), and OpenAICLIP (52.0%). These results demonstrate that CLEAR, with its inherently interpretable design, consistently outperforms standard vision-language foundation models in zero-shot CXR interpretation.

When classifying a CXR using zero-shot transfer with CLEAR, a concept auditing plot (Figure S2) can be generated to visualize the cosine similarity between the CXR and each considered radiological observation. We observed that CLEAR successfully retrieves relevant clinical images for a variety of radiological terms (Figure S3). Concepts with high similarity scores are interpreted by the model as closely matching the ground truth diagnosis, for example, atelectasis (Figure S2a), interstitial and alveolar pattern (Figure S2b), or congestive heart failure (Figure S2c). Notably, CLEAR can also detect semantically meaningful concepts that contribute to prediction errors, even under zero-shot settings, by analyzing visually similar clusters that give rise to significant differences in performance. For instance, in pneumonia detection, test performance differed significantly between VinDr-CXR (Vietnam) and PadChest (Spain), with AUROCs of 0.93 and 0.83, respectively (Figure 2f). Using CLEAR, we identified that the image clusters with the highest error rate within the PadChest test set were those labeled “interstitial lung pattern” and “lingular atelectasis” (Figure 2f and g). The ability of interstitial changes and atelectasis to mask pneumonia-related consolidation on chest imaging is a well-known diagnostic challenge. Our findings align with established radiological principles documented in the Fleischner Society guidelines^67, 68^ which recognize that interstitial patterns create overlapping densities and architectural distortion that can obscure underlying pathology.

### CLEAR improves concept-based image representations for training models

To gain a deeper understanding of the capabilities of our concept-based image embeddings, we trained linear probes in a supervised manner using labels from the MIMIC-CXR training set and evaluated them externally on the labels from the Stanford CheXpert test set. Concept-based image embeddings were first computed by CLEAR and then used as fixed input features for a logistic regression model with a sigmoid activation function to predict the presence or absence of each label (Figure 3a). On CheXpert benchmarks (Figure 3b and Table S5), CLEAR achieved AUROC scores of 87.6%, 90.5%, 72.0%, 88.6%, 93.3%, 94.5%, 83.9%, 89.4%, and 92.8% for atelectasis, consolidation, fracture, lung lesion, lung opacity, pleural effusion, pneumonia, pneumothorax, and support devices, respectively, significantly outperforming the state-of-the-art CheXzero baseline (*P <* 0.01). Overall, CLEAR achieved an average AUROC of 87.0% across all tasks, compared to 71.8% for BiomedCLIP and 67.6% for OpenAICLIP. CLEAR underperformed only on the ”pleural other” class compared with BiomedCLIP (AUROC: 90.8% vs. 91.5%, *P <* 0.01). Thus, CLEAR provides a strong image encoder that performs better than all visual encoders tested, including strong cross-modal self-supervised baselines.

**Figure 3:**
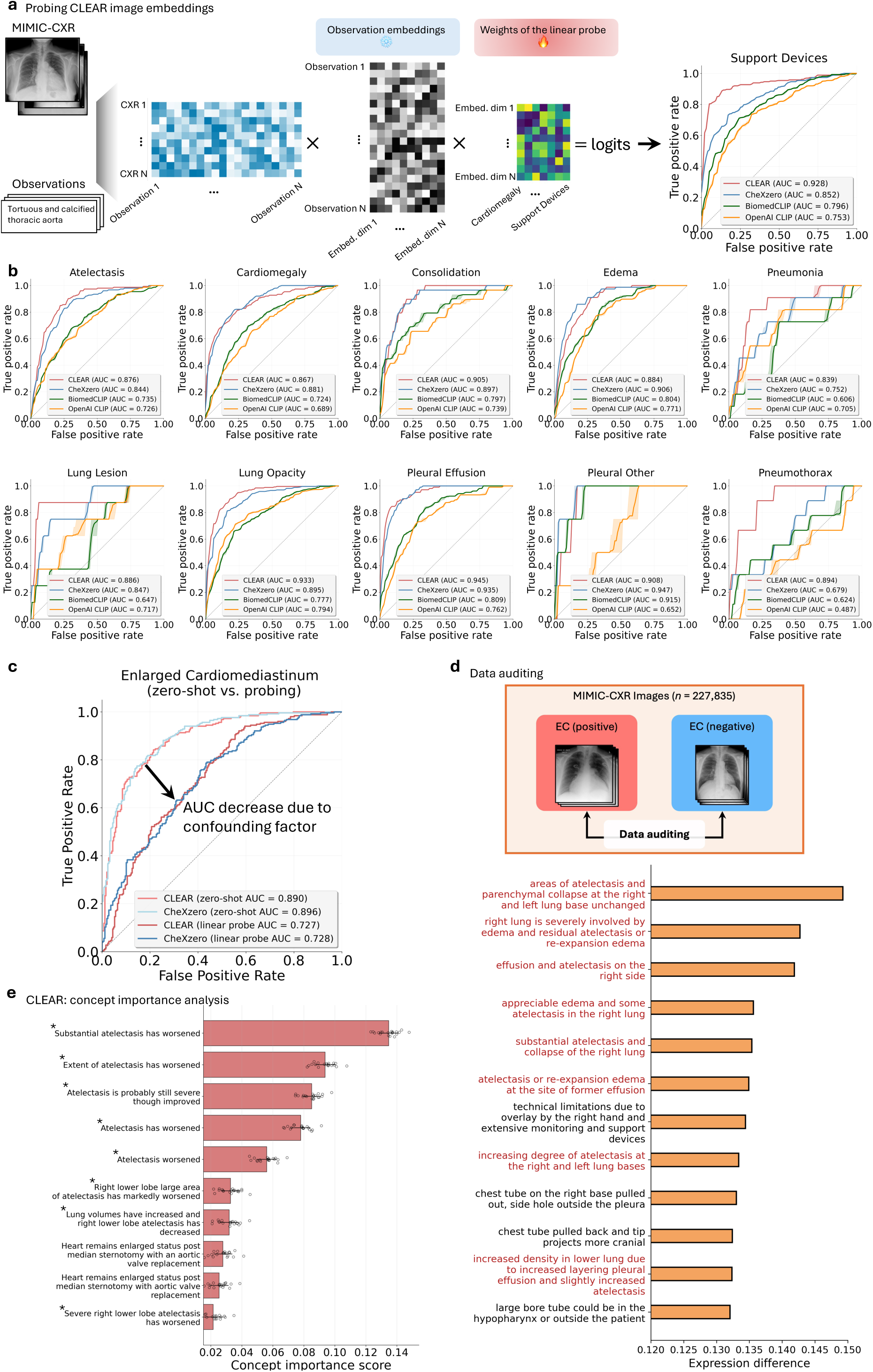
Supervised linear probing experiments and data auditing. **(a)** Linear probing pipeline. CLEAR computes similarities between CXRs and 368,294 radiological observations, projects them via LLM embeddings, and applies learned weights for classification. For the “support devices” example, the AUROC for CLEAR (0.928) is superior to those of CheXzero (0.852), BiomedCLIP (0.796), and OpenAICLIP (0.753). **(b)** ROC curves for supervised classification on CheXpert pathologies. CLEAR (red) achieved AUROCs ranging from 0.839 (pneumonia) to 0.945 (pleural effusion) across 10 diagnostic tasks. **(c)** Performance comparison revealing dataset bias. The enlarged cardiomediastinum example shows AUROC degradation from zero-shot (0.890) to supervised probing (0.727), indicating confounding factors in the training data. **(d)** Data auditing of the MIMIC-CXR training set. Differential concept analysis between positive and negative enlarged cardiomediastinum cases reveals that atelectasis-related features (red text) incorrectly dominate the distinction. **(e)** CLEAR’s concept importance analysis for enlarged cardiomediastinum. Asterisks (∗) mark atelectasis-related concepts that inappropriately influence classification, confirming spurious correlations from label co-occurrence in the dataset.

In addition to making individual-level predictions, linear probes of CLEAR can be used to generate cohort-level bar plots that visualize the contribution of each concept to the predicted class label. This is achieved by multiplying the LLM-generated concept embeddings by the trained weights of the logistic regression model (Figure S4). Concepts with high importance scores are interpreted by the model as closely aligned with the target diagnosis (Figures S5 and S6). For example, Figure S5f shows that the observation “Calcified nodules in the right upper lobe” strongly supports a diagnosis of lung lesion.

However, when examining the concepts contributing to the classification of enlarged cardiomediastinum, we observed that the most influential concepts were in fact related to atelectasis rather than mediastinal enlargement (Figure 3e, concepts marked with ∗). This suggests the presence of a confounding factor: in the MIMIC-CXR dataset, cases labeled with enlarged cardiomediastinum are often co-labeled with atelectasis. The resulting shift in concept attribution led to a noticeable performance drop when transitioning from zero-shot classification (AUROC = 0.890) to linear probing (AUROC = 0.727) for CLEAR and CheXzero, as illustrated in the ROC analysis (Figure 3c). To further investigate this issue, we conducted a data auditing analysis comparing positive and negative cases of enlarged cardiomediastinum from MIMIC-CXR (Figure 3d).^39^ We found that the most distinctive phrases differentiating positive cases also centered around descriptions of atelectasis and effusion, rather than mediastinal contours. This highlights the risk of latent confounding in radiology datasets and underscores the importance of interpretable auditing tools like CLEAR for identifying such biases.

### Expert-level inherently interpretable model building

Linear probing of CLEAR image embeddings enables accurate and interpretable predictions of radiological findings based on a wide range of clinical concepts. However, in some cases, it may be desirable to specialize the model using a high-quality, compact concept set (*n*_C_ *<* 100) to enhance interpretability for human inspection, ideally using as few concepts as possible.^47^ Traditionally, concept lists for standard CBMs are curated through expert guidance or with the assistance of LLMs.^47, 69^ We hypothesized that the optimal concept list for building inherently interpretable models can be derived as a subset of all previously mined clinical observations (Figure S7a). In this section, we present a data-driven, automated approach for selecting a high-quality subset of concepts (*n*_C_ = 68) from our previously constructed concept space (*n*_C_ = 368,294), using the MIMIC-CXR dataset. To construct the CBM concept list, we used linear probes trained on MIMIC-CXR to rank concepts by their importance in classifying 13 diagnostic labels: atelectasis, cardiomegaly, consolidation, edema, enlarged cardiomediastinum, fracture, lung lesion, lung opacity, pleural effusion, pleural other, pneumonia, pneumothorax, and support devices (Figure S7b). We then selected the top ten concepts per label and filtered the resulting set to 68 high-quality concepts using OpenAI’s GPT-4.1 model (Table 1 and Table S6).

**Table 1:** Concepts for building the CBM.

|  |  |
| --- | --- |
| • Lung volumes are lower with bibasilar atelectasis<br>• Lung volumes low on the left due to basilar atelectasis<br>• Severe bibasilar atelectasis<br>• Multiple compression fractures in the lower thoracic and upper lumbar spine<br>• Severe compression fracture of one of the lower thoracic vertebral bodies<br>• Calcified mass in the right upper peripheral lung<br>• Large right upper lobe mass lesion, concerning for lung cancer<br>• Multifocal bilateral opacities and consolidation in the right middle lobe<br>• Large left pleural effusion is a little smaller now than before<br>• Transvenous pacer leads ending in the right atrium, right ventricle, and left ventricle | • Bibasilar atelectasis in the setting of low inspiratory volumes<br>• Lung volumes remain low with right basilar atelectasis<br>• There is now a tiny right apical pneumothorax and a small right lateral basilar pneumothorax, presumably related to interval placement of new chest tube.<br>• Multiple left-sided rib fractures with associated chest wall deformity<br>• Calcified lesion in the left upper lung with associated apical scarring<br>• Right upper lobe cavitary lung mass<br>• Calcified nodule in the left upper lobe<br>• Bilateral pleural effusions have increased, now small-to-moderate on the left and small on the right<br>• Left pacemaker with leads in the lower right atrium and right ventricle<br>• Dual-chamber pacemaker with lead tips in the right atrium and right ventricular apex |

Using the selected concept list, we trained standard CBM baselines in which the model first predicts concept scores from the input using our finetuned vision-language model (i.e., from input to 68 concepts). These predicted concept scores are then used to classify the target labels via a logistic regression model (i.e., from concepts to output, Figure S8b). CLEAR was adapted to use the same 68-concept list for a fair comparison against the CBM baselines (Figure S8c). We evaluated the performance of both inherently interpretable models on the external CheXpert test set, benchmarking them against three board-certified radiologists for the classification of five CheXpert competition pathologies: atelectasis, cardiomegaly, consolidation, edema, and pleural effusion.^20^ We further evaluated model generalizability on three diverse external datasets that differ in geographic origin, patient population, and acquisition protocols: VinDr-CXR, PadChest, and Indiana. We report the widely used Matthews correlation coefficient (MCC) and F1 score (Table 2) in addition to AUROC for each label against ground-truth radiologist annotations (Figure 4).

**Table 2:**
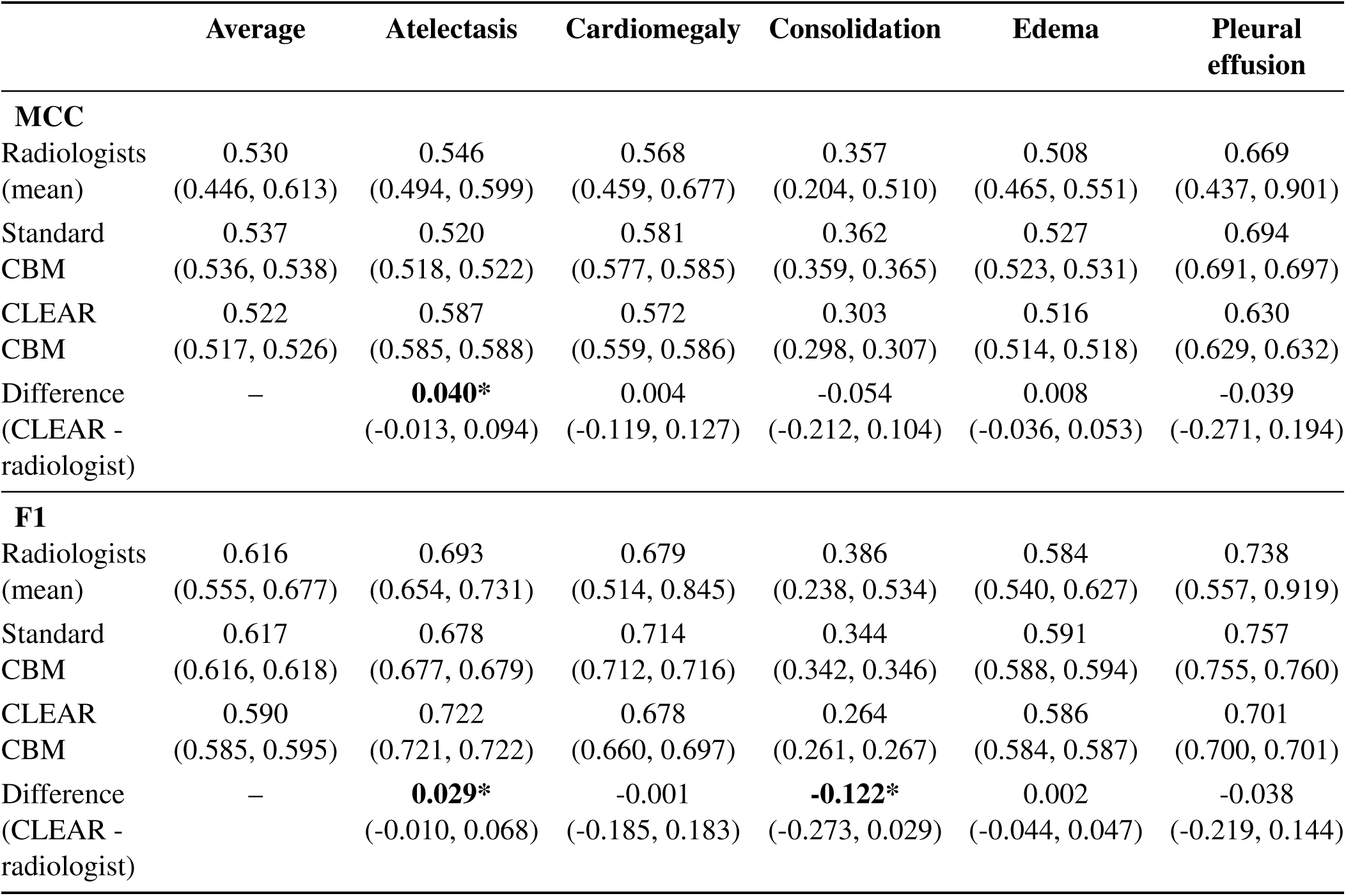
Performance of the CLEAR CBM on the five CheXpert competition pathologies, compared with radiologists and the standard CBM. Values are the mean (95% CI). * indicate *P* < 0.01.

**Figure 4:**
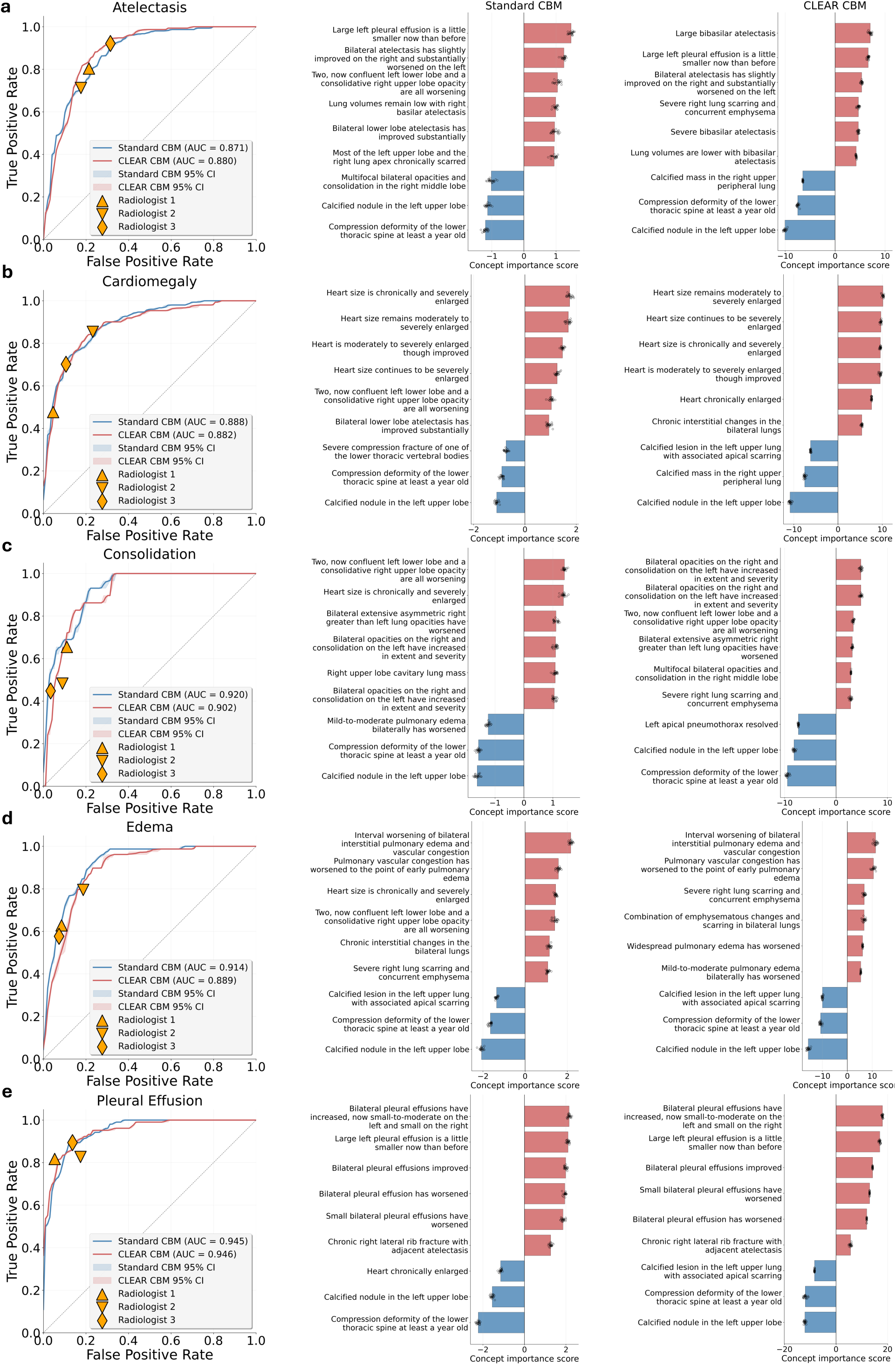
Expert-level concept bottleneck models. **(a)** ROC curves comparing the performance of CLEAR CBM and standard CBM for atelectasis detection against three board-certified radiologists. A model outperforms the radiologists when its ROC curve lies above the radiologists’ operating points (shown as triangles and diamonds). Both models achieve expert-level performance (CLEAR CBM AUROC=0.880, standard CBM AUROC=0.871). Concept importance scores (right panels) reveal that CLEAR CBM correctly prioritizes direct atelectasis indicators like ”Large bibasilar atelectasis” and ”Lung volumes are lower with bibasilar atelectasis,” while standard CBM inappropriately weights ”Large left pleural effusion,” which may confound diagnosis. **(b)** The cardiomegaly detection performance of both CLEAR CBM (AUROC=0.882) and standard CBM (AUROC=0.888) is on par with that of radiologists. Concept analysis shows that CLEAR CBM emphasizes direct cardiac enlargement indicators like “Heart size remains moderately to severely enlarged” while maintaining appropriate weighting of relevant concepts. **(c)** Consolidation detection. Both CBMs achieve high performance (CLEAR CBM AUROC=0.902, standard CBM AUROC=0.920), with concept attributions high-lighting bilateral opacities and lung consolidation patterns as key diagnostic features. **(d)** Edema detection. Standard CBM (AUROC=0.914) slightly outperforms CLEAR CBM (AUROC=0.889). CLEAR CBM correctly emphasizes “Pulmonary vascular congestion” and “Widespread pulmonary edema” while appropriately down-weighting irrelevant findings like “Calcified lesion in the left upper lung.” **(e)** Pleural effusion detection. Both models achieve near-identical expert-level performance (CLEAR CBM AUROC=0.946, Standard CBM AUROC=0.945), with concept scores highlighting bilateral pleural effusion descriptions as primary diagnostic indicators.

On average, across the five pathologies (Table 2), both the standard CBM and the CLEAR CBM demonstrate diagnostic performance comparable to the three board-certified radiologists. In the case of atelectasis, the performance of CLEAR CBM exceeded that of the radiologists, with an MCC of 0.587 (0.546, *P <* 0.01 vs. radiologists; 0.520, *P <* 0.01 vs. standard CBM), and an F1 score of 0.722 (0.693, *P <* 0.01 vs. radiologists; 0.678, *P <* 0.01 vs. standard CBM). By contrast, CLEAR’s F1 score on consolidation (0.264) was significantly lower than that of radiologists (0.386, *P <* 0.01). No statistically significant differences in MCC or F1 score were observed for cardiomegaly, edema, or pleural effusion. Figure 4 shows the ROC curve performance of the CBM models, alongside the radiologists’ operating points. On the external VinDr-CXR, PadChest, and Indiana test sets, CLEAR CBM achieved an AUROC of at least 0.900 on cardiomegaly, edema, and pleural effusion and at least 0.800 on all radiographic findings (Figure S8d).

We further assessed whether the concept attributions generated by CLEAR CBM align with established radiological reasoning (Figure 4, right). For atelectasis (Figure 4a), CLEAR emphasizes hallmark radiographic signs of volume loss used to detect segmental or lobar collapse, including “Large bibasilar atelectasis,” “Lung volumes are lower bilaterally,” and “Severe bibasilar atelectasis”. By contrast, the standard CBM assigns high importance to less specific or potentially confounding features, such as “Large left pleural effusion,” that may mimic or mask atelectasis but are not definitive diagnostic markers. For pulmonary edema (Figure 4d), CLEAR highlights characteristic imaging patterns such as “Pulmonary vascular congestion,” “Widespread pulmonary edema,” and “Mild-to-moderate pulmonary edema bilaterally,” capturing the progression from interstitial to alveolar fluid accumulation. It also appropriately down-weights irrelevant findings, such as “Calcified nodule in the left upper lobe”. By contrast, the standard CBM places greater emphasis on less specific features that may co-occur with edema but lack direct diagnostic value, like “Heart is enlarged”.

## Discussion

Most medical image analysis tools attempt to extract meaningful patterns and discriminative signals from radiological images through end-to-end ML approaches, overlooking the wealth of information contained in radiological reports.^70, 71, 72, 73, 74^ This vast corpus of clinical observations and interpretations allows radiologists to generalize from a few training cases to the presentations encountered in clinical practice, where findings often manifest with substantial variability. In the present study, we leverage the largest publicly available radiology-specific dataset of paired CXRs and reports, comprising around 230,000 studies collected between 2011 and 2016, for large-scale clinical concept mining and the development of a high-performance, auditable foundation model (CLEAR) capable of supporting the detection of a wide range of radiological findings. We generate concept-based image embeddings by projecting all 368,294 concepts into modern LLM semantic spaces. Concept-level auditing is a key advance of CLEAR: for each prediction, the model can surface the clinical observations most influential to its decision, thereby enabling radiologists to inspect, critique, and contextualize model outputs in a manner that aligns with established diagnostic reasoning.

CLEAR’s concept-based embeddings exhibit strong zero-shot recognition capabilities out of the box, frequently outperforming state-of-the-art vision-language foundation models on external benchmarks while preserving interpretability through concept-level auditing. Several recent studies have attempted to leverage image-text pairs from medical or general databases to build vision-language foundation models for biomedicine. Notable among these are CheXzero, BiomedCLIP, and OpenAI CLIP. While CheXzero and BiomedCLIP focus on domain-specific corpora (CXR reports or biomedical literature and images) and OpenAI CLIP scales training to massive, general-domain datasets, these approaches remain fundamentally end-to-end. Their embeddings are not explicitly aligned with radiologist-observed concepts, limiting interpretability and leaving the decision-making process opaque.

Our experiments in concept attribution analysis revealed important insights into clinically similar yet diagnostically distinct cases, as well as key confounding factors. For instance, we found that atelectasis-related concepts dominated the classification of enlarged cardiomediastinum in the established MIMIC-CXR dataset, highlighting how latent label co-occurrences can lead models to learn spurious correlations rather than genuine diagnostic features. This finding underscores the critical importance of interpretable auditing tools in medical AI development, as such biases would remain hidden in black-box models and could potentially propagate to clinical deployment. The ability of CLEAR to identify both spurious dataset correlations and genuine clinical confounders is crucial for building trust in AI systems. Looking forward, these findings suggest several next steps: systematically applying CLEAR to other large-scale CXR datasets to uncover hidden biases, developing quantitative benchmarks for evaluating confounder sensitivity, and integrating auditing pipelines into clinical AI workflows. This extends previous work on dataset auditing and concept bottleneck models by demonstrating that concept attributions generated by CLEAR align with radiological reasoning rather than spurious co-occurrences. In doing so, CLEAR ensures complete interpretability, as clinicians can trace predictions through concept activations with learned weights directly quantifying each concept’s contribution, thereby offering a principled path toward clinically reliable foundation models.

Beyond identifying dataset-specific biases, CLEAR recognizes genuine clinical diagnostic challenges documented in the radiological literature. Chest radiography is widely used as a first-line imaging modality, but its interpretation is notoriously challenging due to overlapping structures, variable image quality, and subtle findings; this complexity was one reason we chose CXR as the focus of this study. Our analysis revealed that pneumonia detection errors were highest in cases with interstitial lung patterns and lingular atelectasis. These findings align with Fleischner Society^67, 68^ and ATS/ERS/JRS/ALAT guidelines,^75^ which recognize these patterns as creating overlapping densities that obscure pneumonia-related consolidation. Beyond pneumonia, CLEAR could next be applied to other difficult scenarios such as differentiating cardiogenic pulmonary edema from bilateral pneumonia, detecting small pneumothoraces hidden at the lung apex, or distinguishing early interstitial lung disease from age-related fibrotic changes.

We additionally investigated the compatibility of our concept-based approach with traditional CBMs for building inherently interpretable models. One limitation of CBMs is that their performance depends heavily on the quality and comprehensiveness of the selected concepts, and prior work has shown that incomplete or poorly aligned concept sets can lead to systematic blind spots and reduced clinical validity.^76, 77, 78, 79^ We found that using a data-driven approach to select high-quality concept subsets (68 concepts) from our large-scale mined observations could yield CBMs that achieve expert-level performance on standardized benchmarks. For the CheXpert competition pathologies, where substantial efforts have been made to build large, annotated datasets suitable for end-to-end supervised learning,^56^ we demonstrated that both standard and CLEAR-based CBMs achieve performance comparable to board-certified radiologists while maintaining full interpretability through compact concept sets. This suggests that the trade-off between performance and interpretability, long considered inevitable in medical AI, may be surmountable through careful concept selection and representation learning. In addition, because CLEAR can identify compact, transferable concept sets without relying on extensive manual labeling, this framework may be particularly valuable in resource-limited settings where large annotated datasets are not available.

A limitation of our study is that our concept space, while comprehensive, is derived from publicly available datasets that may not fully capture the diversity of radiological findings encountered globally or in specialized clinical settings.^80, 81^ While we demonstrated superior performance on multiple external benchmarks, we did not exhaustively investigate robustness across all possible variations in imaging protocols, patient populations, or clinical contexts.^82, 83, 84, 85, 86^ Another consideration is that while CLEAR provides concept-level interpretability, the quality and clinical relevance of these explanations depend on the quality of the original radiological observations extracted from reports. Variations in reporting styles, terminology, and completeness across different institutions and radiologists may influence the concept space learned by the model. In addition, our results showed that different LLM embedding models (e.g., OpenAI, BioMedBERT, Mistral-based embeddings) yielded measurable differences in concept alignment and downstream performance (Figure S1c). This variability underscores that the choice of embedding backbone is itself a limitation, as it may bias which concepts are emphasized or attenuated, and highlights the need for systematic evaluation of embedding models in clinical CBM pipelines. Future work will focus on curating a multi-country, multi-language image-report cohort, enabling broader generalizability and more diverse concept coverage.^87, 88^

## Methods

### Concept space curation

We curated radiological concepts from the MIMIC-CXR dataset, which contains 377,110 chest radiographs corresponding to 227,835 radiographic studies, each accompanied by free-text radiology reports. The curation process presented two primary challenges: (1) extracting discrete clinical observations from narrative radiology reports with variable formatting, abbreviations, and complex medical terminology, and (2) ensuring comprehensive coverage of radiological findings while avoiding redundancy. Given the scale of 227,835 studies, manual extraction was infeasible. We addressed these challenges through a three-step process: (1) automated extraction of observations from report findings and impressions sections using large language models; (2) deduplication and normalization of extracted concepts; and (3) embedding generation using multiple state-of-the-art language models.

For observation extraction, we employed Ministral-8B-Instruct-2410 LLM with few-shot prompting to parse radiology reports and extract discrete clinical findings. We provided the model with exemplar reports demonstrating extraction of observations such as ”mild pulmonary edema” and ”bilateral pleural effusions” to ensure consistent parsing. The model generated structured JSON outputs containing lists of observations for each report. Through this process, we extracted 368,294 unique radiological observations after deduplication and normalization to lowercase.

To generate semantic embeddings, we processed each observation through multiple embedding models, including OpenAI’s text-embedding-3-small (1,536 dimensions), Alibaba Cloud’s Qwen3-Embedding-8B (4,096 dimensions), Salesforce’s SFR-Embedding-Mistral (4,096 dimensions), and Microsoft’s BiomedBERT (768 dimensions). For computational efficiency, we implemented batch processing with sizes up to 32 observations and optional PCA dimension reduction to 768 dimensions while retaining over 95% of explained variance. We found that embeddings generated using Salesforce’s SFR-Embedding-Mistral performed best on downstream validation datasets, and therefore SFR-Embedding-Mistral was used in our CLEAR model. SFR-Embedding-Mistral employs last-token pooling on instruction-formatted inputs, where each observation is prefixed with a task instruction to generate semantic embeddings optimized for medical concept representation.

### Visual-language pretraining

For visual-language pretraining, we employed a contrastive learning framework following the CLIP architecture to align CXR representations with their corresponding radiology reports. The model consisted of an image encoder *f* (·; *θ*) and a text encoder *g*(·; *ϕ*) that project images and text into a shared 768-dimensional embedding space. The image encoder utilized a DINOv2 ViT-B/14 with registers backbone pretrained on 142M natural images,^89^ which processed CXRs at 448×448 pixel resolution through 12 transformer layers with patch size 14×14 and 768-dimensional embeddings. DINOv2 employs a [CLS] token for global image representation alongside register tokens that provide auxiliary storage for processing complex visual patterns without interfering with patch-specific representations. The text encoder employed a 12-layer transformer with 512-dimensional width, 8 attention heads, and 77 maximum token length, following the standard GPT-2 architecture with causal attention masking.

During training, a mini-batch consisted of *M* image-text pairs 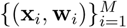, where **x***_i_* denotes the *i*-th CXR and **w***_i_* represents the corresponding radiology report text. Text inputs were tokenized with special tokens, prepending `<|startoftext|>` and appending `<|endoftext|>` tokens. For each pair (**x***_i_,* **w***_i_*), we computed normalized image features **u***_i_* = *f* (**x***_i_*; *θ*)*/*||*f* (**x***_i_*; *θ*)||_2_ and text features **v***_i_* = *g*(**w***_i_*; *ϕ*)*/*||*g*(**w***_i_*; *ϕ*)||_2_, where image features were extracted from the [CLS] token and text features from the `<|endoftext|>` token position. The training objective combined symmetric image-to-text and text-to-image contrastive losses:^65^

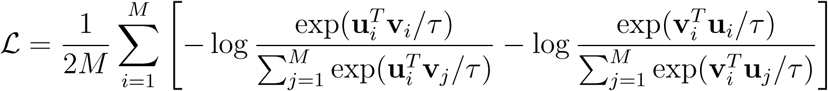

where *τ* denotes a learnable temperature parameter initialized at 0.07. The first term represents the image-to-text contrastive loss, maximizing similarity between paired image and text embeddings relative to negative pairings within the batch. The second term symmetrically computes the text-to-image loss.

Pretraining experiments were conducted on a combined dataset of 873,342 image-text pairs (239,091 patients) aggregated from MIMIC-CXR (377,110 pairs), CheXpert-Plus (223,228 pairs), and ReXGradient (273,004 pairs). Images were resized to 448×448 pixels using bicubic interpolation. Text preprocessing extracted IMPRESSION sections from radiology reports, with exclusion criteria applied to remove reports containing references to prior examinations. Training was distributed across 3 NVIDIA A6000 48GB GPUs with a per-GPU batch size of 64 and gradient accumulation steps of 4, yielding an effective global batch size of 768. Optimization employed AdamW with learning rate 1e-4, weight decay 0.2, and cosine annealing with 500 linear warmup steps over 40 epochs.^90^

### Auditable zero-shot transfer on CXRs

To perform zero-shot classification with CLEAR, we first computed pairwise similarity scores between each input CXR and all 368,294 radiological observations in our concept space. For a given CXR image **x**, we extracted its visual features **I***_e_* = *f* (**x**; *θ*) using the DINOv2 vision encoder after visual-language pretraining and normalized them to unit length. Similarly, each radiological observation **c***_i_* was encoded as **T***_e,i_* = *g*(**c***_i_*; *ϕ*) using the trained text encoder and normalized. The concept similarity vector **s** ∈ ℝ^368,294^ was computed as 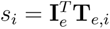, representing the degree to which each concept was present in the image.

To project this concept-based representation into an LLM embedding space for downstream classification, we multiplied the concept similarity vector by a matrix of LLM-generated concept embeddings:

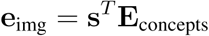

where **E**_concepts_ ∈ ℝ^368,294×^*^d^* contains the *d*-dimensional LLM embeddings for all concepts (e.g., *d* = 4, 096 for SFR-Embedding-Mistral). The resulting image embedding **e**_img_ was normalized to unit length.

To evaluate the zero-shot performance on multi-label classification tasks, we employed a positive-negative softmax evaluation procedure for each disease independently. In contrast to standard CLIP evaluation that normalizes across all disease classes, our approach normalizes between positive and negative versions of the same disease, which is more appropriate for CXR interpretation, where multiple pathologies may coexist in a single image.

For each target disease, we constructed paired prompts: a positive prompt (e.g., ”atelectasis”) and a negative prompt (e.g., ”no atelectasis”), which were encoded using the same LLM embedding model to obtain **e**_pos_ and **e**_neg_. The classification logits were computed as cosine similarities: 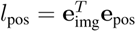 and 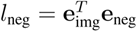. We then applied softmax normalization between these paired logits to obtain the probability of disease presence:

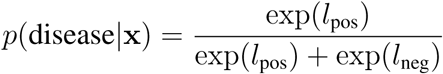

This positive-negative softmax approach treats each disease classification as an independent binary decision, allowing the model to predict multiple concurrent pathologies while maintaining interpretability through the concept similarity scores **s**, which could be visualized to understand which radiological observations contributed most to each prediction.^20^

### Model auditing for zero-shot predictions

To automatically detect imaging characteristics that lead to model errors, we adapted the MONET methodology for auditing our zero-shot predictions.^39^ We sorted test set images into groups based on visual similarity using k-means clustering with 50 principal components of embeddings from the penultimate layer of an ImageNet-pretrained EfficientNetV2-S model.^91^ For each cluster, we calculated disease-specific AUROC scores; clusters with AUROC scores lower than the overall disease AUROC were identified as low-performing clusters. Each low-performing cluster was paired with its most visually similar high-performing counterpart (the cluster with the nearest centroid in Euclidean distance) to understand what differentiates them. We compared mean concept presence scores between paired clusters by computing the mean concept similarity vectors 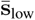 and 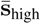, where the difference 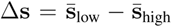 revealed concepts overrepresented in challenging cases. We then ranked the remaining concepts by their differential activation.

### Supervised classification experiments

We performed supervised classification experiments on the 13 CheXpert competition pathologies with labeled training examples from MIMIC-CXR. We used the MIMIC-CXR training set (368,960 images, 64,586 patients) for training and evaluated on the CheXpert test set (500 studies, 500 patients). We considered two approaches: concept-based linear probing using our learned radiological concept representations, and baseline linear probing using frozen visual features from pretrained encoders.

For concept-based experiments, we used the same preprocessing and feature extraction pipeline as our zero-shot classification. Given an input CXR standardized to 448×448 pixels, we extracted visual features **I***_e_* using our trained image encoder with registers and computed similarities with all 368,294 radiological concepts. The resulting concept similarity vector **s** was projected into the SFR-Embedding-Mistral space via matrix multiplication **e**_img_ = **s***^T^* **E**_concepts_, yielding 4,096-dimensional normalized embeddings. For baseline comparison, we considered three visual encoders: (1) CheXzero, which uses ViT-B/32 architecture and was pretrained on MIMIC-CXR using contrastive learning between CXRs and radiology reports, (2) BiomedCLIP, a biomedical vision-language foundation model pretrained on PMC-15M, a dataset of 15 million figure-caption pairs extracted from biomedical research articles in PubMed Central,^64^ and (3) OpenAICLIP with ViT-B/16 architecture, pretrained on 400 million general-domain image-text pairs from the internet. All models used standardized 224×224 input resolution for fair comparison, with model-specific normalization applied during preprocessing.

We trained logistic regression classifiers on top of the frozen feature representations using binary cross-entropy loss. Training employed the Adam optimizer with learning rate 2 × 10^−4^ and no weight decay. We used a batch size of 512 and trained for a maximum of 200 epochs with early stopping based on validation AUROC (patience of 10 epochs). To ensure robust performance estimates, we repeated all experiments with 20 different random seeds.

For interpretability analysis, we examined the learned linear weights to identify which concepts were most strongly associated with each label. Given that our logistic regression model operates on LLM-projected features (**e**_img_ = **s***^T^* **E**_concepts_), the learned weight vector **w***_d_* ∈ ℝ^4,096^ for disease *d* represents importance weights in the LLM embedding space. To interpret which radiological concepts contribute most to each disease prediction, we computed the alignment between **w***_d_* and each concept’s LLM embedding vector from **E**_concepts_ using cosine similarity. Concepts with high positive alignment scores indicate radiological findings that increase the predicted logit of disease presence, while those with negative alignment suppress it.

### Data auditing for supervised experiments

CLEAR’s ability to map images and texts onto the co-embedding space enabled us to describe the different characteristics between two sets of images in natural language through concept differential analysis. This technique revealed spurious correlations in training data that might lead models to learn shortcuts rather than genuine diagnostic features.

Following MONET, for each disease in the MIMIC-CXR training set, we denoted the disease-positive images as 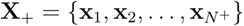 and disease-negative images as 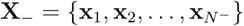, where we sampled *N*^+^ = *N*^−^ = 1, 000 images from each set.^39^ Given our list of radiological concepts 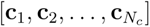 to investigate, we first obtained the prototype embedding of each image set by computing an average of normalized image embeddings: 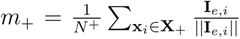 and 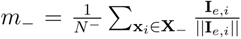, where **I***_e,i_* = *f* (**x***_i_*; *ϕ*) is the image embedding for image **x***_i_*. We then calculated the displacement vector from *m*_−_ to *m*_+_ by subtracting out the two prototype embeddings: *m*_Δ_ = *m*_+_ − *m*_−_. Finally, we obtained a differential concept expression score by computing the dot product between the displacement vector and normalized embeddings of concept prompts: 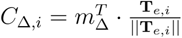, where **T***_e,i_* = *g*(**c***_i_*; *ϕ*) is the text embedding for concept **c***_i_*. This score measures how much more each concept is differentially expressed in **X**_+_ than in **X**_−_.

### Concept bottleneck models

CBMs achieve interpretability by constraining predictions to flow through a human-understandable intermediate representation, forcing the model to first predict the presence of predefined concepts before making final disease classifications. To build CBMs that provide clinically meaningful explanations while maintaining high predictive performance, we systematically curated a compact concept set from our comprehensive space of 368,294 radiological observations. Our data-driven selection process began by training linear probes on concept-based image embeddings from MIMIC-CXR to identify which concepts were most predictive of each disease. For each of the 13 diagnostic labels, we analyzed the learned weight vector **w***_d_* ∈ ℝ^4,096^ by computing its cosine similarity with every concept’s LLM embedding. This analysis revealed which radiological observations aligned most strongly with each disease’s decision boundary. We selected the top 10 positively-aligned concepts per label, yielding approximately 130 candidate concepts that demonstrated strong predictive signal across all diseases. To ensure clinical coherence and remove redundancies, we employed OpenAI’s GPT-4.1 to filter this candidate set, resulting in 68 high-quality diagnostic concepts that span key radiological findings.

The standard CBM architecture implements the bottleneck layer **z***_c_* : **x**_image_ → ℝ^68^ as a direct projection of visual features onto the selected concept space. For an input CXR **x**, we compute concept activations as 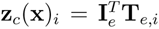, where **I***_e_* = *f* (**x**; *θ*) is the visual feature from our trained DINOv2 encoder and **T***_e,i_* is the *i*-th concept’s text embedding. These continuous-valued activations quantify each concept’s presence in the image without requiring binarization. Final disease predictions employ linear classifiers on the 68-dimensional concept activation vector: 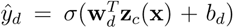 for disease *d*. This ensures complete interpretability as clinicians can trace predictions through concept activations, with learned weights **w***_d_* directly quantifying each concept’s contribution to the disease prediction.

### Evaluation metrics

For classification tasks, we employed the area under the receiver operating characteristic curve (AUROC) as our primary evaluation metric, calculated from the true positive rate against the false positive rate as the classification threshold varies. For multi-label CXR interpretation, we computed AUROC independently for each pathology and reported both individual and macro-averaged values. When comparing with radiologist performance on CheXpert competition pathologies, we additionally reported the Matthews correlation coefficient (MCC) and F1 score. The MCC provides a balanced measure accounting for all confusion matrix categories (ranging from −1 to +1), and the F1 score represents the harmonic mean of precision and recall.

### Evaluation datasets

VinDr-CXR is a publicly available Vietnamese CXR dataset comprising 18,000 frontal radiographs collected from two major hospitals in Vietnam between 2018 and 2020.^92^ We evaluated our models on the official test set of 3,000 CXRs, which includes consensus annotations for 21 radiological findings provided by three board-certified radiologists. This dataset served as a benchmark for evaluating both zero-shot disease classification and the performance of our CBMs.

PadChest is a large-scale Spanish CXR dataset comprising 160,868 images collected from 67,625 patients, collected at San Juan Hospital between 2009 and 2017.^93^ For evaluation, we followed the protocol of Han et al.^43^ to curate a subset of 7,943 frontal radiographs from 7,272 patients, each annotated with physician-verified labels for 55 phenotype-relevant findings. This dataset served as a benchmark for assessing both zero-shot disease classification and the performance of our CBMs.

The Indiana University Chest X-ray Collection (Indiana CXR) comprises 7,470 chest radiographs from Indiana University hospitals, each paired with its corresponding radiology report.^94^ Following preprocessing and label extraction, we obtained 7,466 frontal CXRs annotated for 30 radiological findings. This dataset was used as a benchmark to evaluate both zero-shot disease classification and the performance of our CBMs.

The CheXpert test set is the official held-out test set from the Stanford CheXpert competition and contains 500 chest radiographs from 500 unique patients collected at Stanford Hospital.^56^ Each image was labeled by five board-certified radiologists for the 14 competition pathologies: atelectasis, cardiomegaly, consolidation, edema, enlarged cardiomediastinum, fracture, lung lesion, lung opacity, no finding, pleural effusion, pleural other, pneumonia, pneumothorax, and support devices. We used this dataset for supervised classification experiments and for benchmarking CBM performance against three board-certified radiologists on five selected pathologies (atelectasis, cardiomegaly, consolidation, edema, and pleural effusion) where ground-truth consensus labels and individual radiologist annotations were available.

### Statistical analysis

Statistical significance of performance differences between models was assessed using two-sided paired permutation tests with 10,000 permutations, where predictions from compared models were randomly swapped to generate a null distribution. The *P* value was calculated as the proportion of permuted differences exceeding the observed difference in absolute value, with *P <* 0.01 considered statistically significant. For performance evaluation, we computed 95% confidence intervals using nonparametric bootstrapping with 1,000 resamples drawn with replacement from the test set. For CBM experiments comparing with radiologist performance, we additionally used the t-distribution to compute confidence intervals to account for the small sample size of radiologists. To ensure robust performance estimates in supervised experiments and CBM, we employed 20 different random seeds for model training and aggregated the results across seeds, computing mean and standard deviation of performance metrics.

## Data availability

The MIMIC-CXR dataset is publicly available from PhysioNet at https://physionet.org/content/mimic-cxr/2.1.0/. The CheXpert-Plus dataset can be accessed at https://aimi.stanford.edu/datasets/chexpert-plus. The ReXGradient data can be accessed at https://huggingface.co/datasets/rajpurkarlab/ReXGradient-160K. The PadChest dataset is available from https://bimcv.cipf.es/bimcv-projects/padchest. The VinDr-CXR dataset can be obtained from https://vindr.ai/cxr. The Indiana University Chest X-ray Collection is accessible through https://openi.nlm.nih.gov/. The CheXpert test set requires official competition registration at https://github.com/rajpurkarlab/cheXpert-test-set-labels.

## Code availability

Model weights for CLEAR can be accessed for academic research purposes at http://huggingface.co/[to-be-released]. Code for using the pretrained model is provided at https://github.com/peterhan91/CLEAR. We have documented all technical deep learning methods and software libraries used in the study while ensuring the paper is accessible to the broader clinical and scientific audience.

## Ethics Statement

This study utilized only publicly available, de-identified datasets that have received appropriate institutional review board approvals from their respective institutions. No additional patient data were collected for this research. The MIMIC-CXR, CheXpert-Plus, ReXGradient, PadChest, VinDr-CXR, and Indiana University datasets were all previously de-identified in compliance with the Health Insurance Portability and Accountability Act (HIPAA) Safe Harbor provision. As this study involved only secondary analysis of existing, publicly available data, no additional ethics approval was required.

## Author Contributions

T.H., R.W., C.D., L.S. and D.T. conceived the study. T.H. and D.T. designed and implemented the CLEAR architecture. T.H. performed the concept extraction and embedding experiments. T.H. and R.W. conducted the visual-language pretraining. T.H. and R.W. performed the zero-shot evaluation experiments. T.H. implemented and evaluated the concept bottleneck models. D.T. and E.M.B. provided clinical insights and evaluated model interpretability. T.H. and D.T. analyzed the results. T.H. and D.T. wrote the manuscript with input from all authors.

## Supplementary Information

**Table S1:** Zero-shot performance on the VinDr-CXR dataset. AUROC values with 95% confidence intervals. Bold indicates best performance.

| Finding | N | CLEAR | CheXzero | BiomedCLIP | OpenAICLIP |
| --- | --- | --- | --- | --- | --- |
| Pneumonia | 246 | **0.937 (0.922, 0.950)** | 0.889 (0.865, 0.912) | 0.675 (0.636, 0.713) | 0.545 (0.507, 0.583) |
| Pleural effusion | 111 | 0.904 (0.882, 0.923) | **0.959 (0.934, 0.979)** | 0.871 (0.833, 0.908) | 0.529 (0.469, 0.587) |
| Lung tumor | 80 | **0.884 (0.847, 0.918)** | 0.810 (0.757, 0.860) | 0.784 (0.731, 0.833) | 0.541 (0.480, 0.598) |
| Pneumothorax | 18 | **0.879 (0.837, 0.918)** | 0.828 (0.744, 0.898) | 0.662 (0.487, 0.822) | 0.450 (0.342, 0.571) |
| Mediastinal shift | 20 | **0.878 (0.825, 0.923)** | 0.846 (0.750, 0.919) | 0.828 (0.720, 0.918) | 0.485 (0.354, 0.606) |
| Consolidation | 96 | 0.877 (0.855, 0.896) | **0.905 (0.869, 0.939)** | 0.615 (0.552, 0.679) | 0.422 (0.365, 0.478) |
| ILD | 221 | 0.816 (0.789, 0.840) | **0.838 (0.811, 0.866)** | 0.711 (0.675, 0.749) | 0.603 (0.565, 0.640) |
| Lung Opacity | 84 | 0.809 (0.768, 0.845) | **0.812 (0.768, 0.851)** | 0.679 (0.619, 0.736) | 0.558 (0.488, 0.625) |
| Nodule/Mass | 176 | **0.793 (0.754, 0.829)** | 0.662 (0.622, 0.700) | 0.536 (0.490, 0.583) | 0.565 (0.521, 0.607) |
| Atelectasis | 86 | **0.785 (0.735, 0.834)** | 0.745 (0.696, 0.789) | 0.709 (0.649, 0.769) | 0.554 (0.496, 0.615) |
| Cardiomegaly | 309 | 0.782 (0.753, 0.810) | **0.936 (0.924, 0.948)** | 0.769 (0.740, 0.796) | 0.591 (0.557, 0.623) |
| Rib fracture | 11 | **0.776 (0.659, 0.873)** | 0.579 (0.394, 0.767) | 0.511 (0.327, 0.683) | 0.491 (0.325, 0.643) |
| Pulmonary fibrosis | 217 | **0.768 (0.735, 0.800)** | 0.668 (0.631, 0.706) | 0.657 (0.617, 0.696) | 0.604 (0.568, 0.643) |
| Other lesion | 94 | **0.763 (0.720, 0.808)** | 0.745 (0.695, 0.793) | 0.560 (0.499, 0.624) | 0.571 (0.517, 0.625) |
| Infiltration | 58 | 0.725 (0.652, 0.791) | **0.843 (0.780, 0.894)** | 0.451 (0.373, 0.534) | 0.519 (0.437, 0.600) |
| Pleural thickening | 169 | **0.722 (0.677, 0.765)** | 0.688 (0.648, 0.728) | 0.635 (0.588, 0.681) | 0.608 (0.564, 0.651) |
| Aortic enlargement | 220 | 0.709 (0.673, 0.745) | **0.758 (0.726, 0.788)** | 0.726 (0.690, 0.761) | 0.543 (0.506, 0.579) |
| No finding | 2051 | **0.693 (0.672, 0.714)** | 0.139 (0.125, 0.154) | 0.369 (0.345, 0.390) | 0.506 (0.484, 0.528) |
| Tuberculosis | 164 | 0.666 (0.623, 0.710) | **0.731 (0.690, 0.770)** | 0.647 (0.600, 0.692) | 0.502 (0.460, 0.546) |
| Other disease | 657 | **0.636 (0.616, 0.656)** | 0.604 (0.579, 0.627) | 0.455 (0.430, 0.481) | 0.489 (0.464, 0.514) |
| Calcification | 194 | 0.617 (0.573, 0.658) | **0.760 (0.728, 0.791)** | 0.604 (0.564, 0.644) | 0.382 (0.336, 0.424) |

**Table S2:** Zero-shot performance on the PadChest dataset. AUROC values with 95% confidence intervals. Bold indicates best performance.

| Finding | N | CLEAR | CheXzero | BiomedCLIP | OpenAICLIP |
| --- | --- | --- | --- | --- | --- |
| pleural effusion | 376 | **0.966 (0.958, 0.972)** | 0.965 (0.955, 0.974) | 0.928 (0.915, 0.942) | 0.565 (0.538, 0.592) |
| pulmonary mass | 42 | **0.941 (0.907, 0.968)** | 0.883 (0.820, 0.934) | 0.888 (0.826, 0.943) | 0.567 (0.484, 0.646) |
| consolidation | 63 | **0.916 (0.893, 0.936)** | 0.875 (0.823, 0.921) | 0.757 (0.698, 0.815) | 0.530 (0.452, 0.606) |
| heart insufficiency | 95 | 0.915 (0.896, 0.931) | **0.924 (0.906, 0.939)** | 0.789 (0.748, 0.833) | 0.349 (0.296, 0.403) |
| hilar congestion | 117 | 0.845 (0.820, 0.869) | **0.888 (0.867, 0.909)** | 0.725 (0.682, 0.769) | 0.576 (0.524, 0.629) |
| hypoexpansion | 27 | **0.842 (0.792, 0.885)** | 0.764 (0.670, 0.852) | 0.634 (0.530, 0.742) | 0.450 (0.346, 0.556) |
| interstitial pattern | 368 | **0.841 (0.822, 0.860)** | 0.819 (0.795, 0.839) | 0.745 (0.719, 0.772) | 0.424 (0.394, 0.456) |
| laminar atelectasis | 235 | **0.840 (0.814, 0.865)** | 0.685 (0.650, 0.716) | 0.500 (0.462, 0.539) | 0.515 (0.479, 0.549) |
| pneumonia | 336 | **0.835 (0.811, 0.856)** | 0.816 (0.792, 0.840) | 0.645 (0.614, 0.674) | 0.498 (0.465, 0.530) |
| cardiomegaly | 739 | 0.825 (0.813, 0.835) | **0.896 (0.887, 0.905)** | 0.773 (0.758, 0.788) | 0.499 (0.475, 0.521) |
| atelectasis | 145 | **0.820 (0.782, 0.853)** | 0.786 (0.748, 0.820) | 0.561 (0.508, 0.621) | 0.434 (0.387, 0.481) |
| lobar atelectasis | 35 | 0.818 (0.760, 0.874) | 0.721 (0.634, 0.803) | **0.826 (0.743, 0.899)** | 0.534 (0.429, 0.638) |
| infiltrates | 253 | **0.795 (0.767, 0.821)** | 0.750 (0.716, 0.780) | 0.569 (0.528, 0.610) | 0.483 (0.446, 0.519) |
| alveolar pattern | 271 | 0.791 (0.769, 0.813) | **0.907 (0.886, 0.926)** | 0.428 (0.388, 0.467) | 0.448 (0.412, 0.485) |
| ground glass pattern | 31 | **0.779 (0.705, 0.847)** | 0.582 (0.461, 0.698) | 0.625 (0.522, 0.728) | 0.553 (0.455, 0.647) |
| rib fracture | 32 | **0.777 (0.729, 0.820)** | 0.488 (0.378, 0.596) | 0.628 (0.513, 0.742) | 0.354 (0.252, 0.469) |
| bronchiectasis | 122 | **0.762 (0.726, 0.801)** | 0.744 (0.697, 0.786) | 0.658 (0.609, 0.702) | 0.510 (0.459, 0.560) |
| pleural thickening | 47 | 0.762 (0.700, 0.821) | **0.793 (0.732, 0.851)** | 0.657 (0.577, 0.736) | 0.508 (0.426, 0.587) |
| costophrenic angle blunting | 345 | 0.759 (0.734, 0.784) | **0.787 (0.764, 0.809)** | 0.483 (0.450, 0.515) | 0.546 (0.515, 0.577) |
| tuberculosis sequelae | 42 | **0.758 (0.691, 0.817)** | 0.616 (0.528, 0.702) | 0.593 (0.505, 0.678) | 0.523 (0.436, 0.613) |
| hypoexpansion basal | 36 | 0.736 (0.676, 0.794) | **0.753 (0.691, 0.808)** | 0.524 (0.425, 0.620) | 0.484 (0.384, 0.576) |
| hiatal hernia | 115 | 0.734 (0.704, 0.764) | **0.759 (0.724, 0.792)** | 0.489 (0.440, 0.540) | 0.573 (0.523, 0.622) |
| hemidiaphragm elevation | 149 | **0.726 (0.694, 0.755)** | 0.631 (0.579, 0.679) | 0.660 (0.614, 0.701) | 0.528 (0.484, 0.572) |
| aortic button enlargement | 49 | **0.726 (0.666, 0.785)** | 0.716 (0.647, 0.782) | 0.672 (0.599, 0.745) | 0.407 (0.326, 0.492) |
| descendent aortic elongation | 35 | 0.717 (0.666, 0.766) | **0.759 (0.701, 0.811)** | 0.554 (0.457, 0.658) | 0.508 (0.398, 0.625) |
| aortic atheromatosis | 322 | **0.699 (0.676, 0.722)** | 0.567 (0.536, 0.598) | 0.557 (0.524, 0.589) | 0.598 (0.568, 0.629) |
| supra aortic elongation | 28 | 0.694 (0.630, 0.754) | **0.704 (0.630, 0.776)** | 0.564 (0.437, 0.691) | 0.431 (0.320, 0.539) |
| chronic changes | 749 | **0.685 (0.668, 0.703)** | 0.658 (0.640, 0.677) | 0.529 (0.509, 0.550) | 0.561 (0.540, 0.583) |
| aortic elongation | 623 | 0.685 (0.668, 0.700) | **0.692 (0.673, 0.709)** | 0.575 (0.552, 0.599) | 0.547 (0.522, 0.572) |
| nodule | 149 | **0.665 (0.628, 0.705)** | 0.546 (0.499, 0.597) | 0.521 (0.472, 0.571) | 0.542 (0.493, 0.589) |
| osteopenia | 44 | **0.663 (0.583, 0.729)** | 0.657 (0.569, 0.737) | 0.639 (0.569, 0.706) | 0.564 (0.474, 0.655) |
| fibrotic band | 250 | **0.654 (0.622, 0.685)** | 0.575 (0.539, 0.609) | 0.518 (0.482, 0.553) | 0.547 (0.512, 0.583) |
| hyperinflated lung | 36 | 0.650 (0.559, 0.737) | **0.768 (0.683, 0.843)** | 0.728 (0.641, 0.808) | 0.545 (0.460, 0.625) |
| calcified granuloma | 141 | **0.647 (0.602, 0.689)** | 0.513 (0.465, 0.562) | 0.452 (0.407, 0.498) | 0.533 (0.485, 0.583) |
| hilar enlargement | 91 | **0.646 (0.594, 0.695)** | 0.622 (0.565, 0.682) | 0.548 (0.486, 0.605) | 0.482 (0.422, 0.539) |
| vertebral anterior compression | 116 | **0.639 (0.586, 0.690)** | 0.633 (0.583, 0.685) | 0.537 (0.484, 0.590) | 0.581 (0.528, 0.632) |
| kyphosis | 221 | 0.630 (0.598, 0.663) | **0.653 (0.619, 0.686)** | 0.527 (0.487, 0.566) | 0.428 (0.390, 0.464) |
| copd signs | 905 | 0.615 (0.599, 0.633) | **0.752 (0.735, 0.768)** | 0.548 (0.528, 0.566) | 0.584 (0.564, 0.603) |
| calcified densities | 65 | **0.612 (0.552, 0.672)** | 0.559 (0.500, 0.614) | 0.426 (0.350, 0.498) | 0.435 (0.357, 0.511) |
| granuloma | 35 | **0.611 (0.530, 0.691)** | 0.516 (0.436, 0.595) | 0.475 (0.389, 0.566) | 0.565 (0.463, 0.665) |
| mediastinic lipomatosis | 28 | 0.610 (0.541, 0.671) | **0.654 (0.560, 0.745)** | 0.499 (0.383, 0.602) | 0.604 (0.503, 0.702) |

**Table S3:** Zero-shot performance on the PadChest dataset (continued). AUROC values with 95% confidence intervals. Bold indicates best performance.

| Finding | N | CLEAR | CheXzero | BiomedCLIP | OpenAICLIP |
| --- | --- | --- | --- | --- | --- |
| sclerotic bone lesion | 30 | 0.598 (0.487, 0.696) | 0.415 (0.313, 0.510) | 0.556 (0.449, 0.650) | **0.658 (0.574, 0.734)** |
| bullas | 33 | **0.597 (0.501, 0.688)** | 0.459 (0.369, 0.548) | 0.506 (0.433, 0.587) | 0.465 (0.365, 0.566) |
| vascular hilar enlargement | 276 | 0.588 (0.556, 0.619) | **0.625 (0.594, 0.657)** | 0.546 (0.513, 0.580) | 0.542 (0.507, 0.578) |
| pseudonodule | 145 | 0.587 (0.544, 0.634) | 0.468 (0.425, 0.507) | 0.551 (0.502, 0.596) | **0.610 (0.565, 0.653)** |
| diaphragmatic eventration | 76 | **0.586 (0.536, 0.632)** | 0.584 (0.526, 0.639) | 0.423 (0.360, 0.487) | 0.545 (0.479, 0.608) |
| nipple shadow | 76 | 0.576 (0.510, 0.642) | 0.559 (0.500, 0.618) | 0.495 (0.433, 0.554) | **0.585 (0.521, 0.646)** |
| flattened diaphragm | 36 | **0.568 (0.486, 0.649)** | 0.514 (0.435, 0.594) | 0.527 (0.443, 0.605) | 0.433 (0.333, 0.533) |
| scoliosis | 544 | 0.566 (0.543, 0.589) | **0.602 (0.576, 0.627)** | 0.554 (0.528, 0.578) | 0.500 (0.474, 0.523) |
| volume loss | 156 | 0.563 (0.523, 0.602) | **0.618 (0.576, 0.661)** | 0.547 (0.496, 0.598) | 0.577 (0.531, 0.620) |
| apical pleural thickening | 292 | 0.558 (0.524, 0.589) | **0.573 (0.541, 0.604)** | 0.537 (0.502, 0.572) | 0.551 (0.518, 0.580) |
| goiter | 54 | 0.541 (0.473, 0.603) | 0.570 (0.505, 0.638) | **0.616 (0.553, 0.673)** | 0.539 (0.462, 0.616) |
| air trapping | 354 | 0.514 (0.484, 0.545) | 0.470 (0.442, 0.498) | **0.581 (0.549, 0.611)** | 0.564 (0.534, 0.593) |
| gynecomastia | 62 | 0.512 (0.452, 0.568) | 0.494 (0.428, 0.556) | **0.675 (0.608, 0.737)** | 0.641 (0.573, 0.709) |
| azygos lobe | 36 | **0.475 (0.377, 0.568)** | 0.414 (0.327, 0.502) | 0.467 (0.386, 0.545) | 0.469 (0.381, 0.553) |

**Table S4:** Zero-shot performance on the Indiana dataset. AUROC values with 95% confidence intervals. Bold indicates best performance.

| Finding | N | CLEAR | CheXzero | BiomedCLIP | OpenAICLIP |
| --- | --- | --- | --- | --- | --- |
| pleural effusion | 286 | 0.935 (0.920, 0.948) | **0.937 (0.921, 0.951)** | 0.775 (0.748, 0.802) | 0.567 (0.530, 0.605) |
| consolidation | 53 | **0.926 (0.902, 0.948)** | 0.874 (0.818, 0.926) | 0.715 (0.645, 0.785) | 0.485 (0.408, 0.568) |
| pulmonary edema | 87 | 0.887 (0.852, 0.915) | **0.920 (0.885, 0.948)** | 0.725 (0.676, 0.772) | 0.457 (0.392, 0.518) |
| pneumonia | 77 | **0.882 (0.845, 0.915)** | 0.823 (0.774, 0.877) | 0.553 (0.491, 0.613) | 0.532 (0.469, 0.594) |
| pulmonary fibrosis | 34 | **0.881 (0.817, 0.933)** | 0.770 (0.688, 0.849) | 0.655 (0.559, 0.738) | 0.669 (0.568, 0.765) |
| pulmonary congestion | 135 | 0.872 (0.844, 0.897) | **0.892 (0.866, 0.917)** | 0.664 (0.619, 0.707) | 0.525 (0.476, 0.577) |
| airspace disease | 224 | **0.845 (0.821, 0.868)** | 0.729 (0.695, 0.763) | 0.601 (0.567, 0.636) | 0.503 (0.462, 0.544) |
| cardiomegaly | 655 | 0.840 (0.827, 0.853) | **0.902 (0.892, 0.913)** | 0.662 (0.642, 0.681) | 0.485 (0.460, 0.509) |
| infiltrate | 116 | **0.798 (0.759, 0.831)** | 0.733 (0.678, 0.786) | 0.652 (0.599, 0.707) | 0.504 (0.443, 0.562) |
| mass | 32 | **0.794 (0.732, 0.851)** | 0.697 (0.613, 0.775) | 0.657 (0.582, 0.728) | 0.499 (0.406, 0.591) |
| pulmonary atelectasis | 616 | **0.776 (0.758, 0.794)** | 0.756 (0.736, 0.774) | 0.509 (0.483, 0.533) | 0.512 (0.488, 0.536) |
| pulmonary emphysema | 74 | 0.750 (0.680, 0.811) | **0.833 (0.790, 0.871)** | 0.584 (0.523, 0.648) | 0.559 (0.484, 0.628) |
| pneumothorax | 44 | 0.735 (0.658, 0.806) | **0.776 (0.706, 0.841)** | 0.524 (0.435, 0.607) | 0.379 (0.300, 0.465) |
| cicatrix | 355 | **0.729 (0.704, 0.754)** | 0.628 (0.600, 0.655) | 0.533 (0.501, 0.564) | 0.550 (0.520, 0.580) |
| opacity | 840 | 0.717 (0.699, 0.734) | **0.752 (0.735, 0.770)** | 0.517 (0.497, 0.537) | 0.540 (0.521, 0.561) |
| normal | 2695 | **0.681 (0.669, 0.693)** | 0.632 (0.620, 0.644) | 0.530 (0.517, 0.543) | 0.483 (0.469, 0.496) |
| atherosclerosis | 233 | 0.661 (0.630, 0.692) | **0.665 (0.636, 0.696)** | 0.546 (0.507, 0.582) | 0.585 (0.548, 0.623) |
| arthritis | 65 | **0.652 (0.584, 0.718)** | 0.352 (0.292, 0.413) | 0.490 (0.422, 0.556) | 0.500 (0.433, 0.569) |
| emphysema | 116 | 0.647 (0.598, 0.697) | **0.869 (0.846, 0.891)** | 0.482 (0.426, 0.538) | 0.534 (0.476, 0.589) |
| deformity | 210 | **0.641 (0.606, 0.678)** | 0.576 (0.535, 0.614) | 0.546 (0.505, 0.585) | 0.465 (0.426, 0.506) |
| kyphosis | 58 | 0.624 (0.559, 0.689) | **0.704 (0.647, 0.761)** | 0.504 (0.417, 0.585) | 0.355 (0.271, 0.432) |
| calcified granuloma | 510 | **0.605 (0.580, 0.629)** | 0.478 (0.452, 0.502) | 0.495 (0.470, 0.522) | 0.512 (0.486, 0.537) |
| calcinosis | 558 | **0.586 (0.562, 0.608)** | 0.429 (0.406, 0.453) | 0.506 (0.480, 0.532) | 0.511 (0.485, 0.535) |
| nodule | 211 | **0.586 (0.546, 0.625)** | 0.518 (0.480, 0.559) | 0.510 (0.474, 0.546) | 0.561 (0.522, 0.599) |
| granuloma | 103 | **0.577 (0.523, 0.626)** | 0.411 (0.355, 0.466) | 0.521 (0.466, 0.579) | 0.547 (0.493, 0.602) |
| spondylosis | 126 | 0.566 (0.514, 0.619) | 0.439 (0.393, 0.485) | 0.470 (0.420, 0.520) | **0.577 (0.532, 0.625)** |
| osteophyte | 137 | 0.545 (0.502, 0.589) | **0.602 (0.556, 0.647)** | 0.477 (0.422, 0.526) | 0.542 (0.494, 0.591) |
| granulomatous disease | 213 | 0.518 (0.482, 0.551) | **0.555 (0.518, 0.595)** | 0.471 (0.437, 0.505) | 0.498 (0.459, 0.535) |
| scoliosis | 177 | 0.505 (0.458, 0.549) | **0.600 (0.557, 0.642)** | 0.493 (0.450, 0.535) | 0.526 (0.485, 0.568) |
| lucency | 51 | 0.466 (0.391, 0.544) | 0.332 (0.264, 0.408) | **0.528 (0.457, 0.602)** | 0.487 (0.404, 0.578) |

**Table S5:** Supervised performance on the CheXpert dataset. AUROC values with 95% confidence intervals. Bold indicates best performance.

| Finding | CLEAR | CheXzero | BiomedCLIP | OpenAICLIP |
| --- | --- | --- | --- | --- |
| Atelectasis | **0.876 (0.876, 0.876)** | 0.844 (0.844, 0.845) | 0.735 (0.733, 0.737) | 0.726 (0.725, 0.727) |
| Cardiomegaly | 0.867 (0.867, 0.868) | **0.881 (0.881, 0.882)** | 0.724 (0.724, 0.725) | 0.689 (0.688, 0.691) |
| Consolidation | **0.905 (0.905, 0.905)** | 0.897 (0.896, 0.897) | 0.797 (0.795, 0.799) | 0.739 (0.738, 0.739) |
| Edema | 0.884 (0.884, 0.884) | **0.906 (0.905, 0.906)** | 0.804 (0.803, 0.804) | 0.771 (0.770, 0.773) |
| Enlarged Cardiomedastinum | **0.727 (0.727, 0.728)** | 0.727 (0.726, 0.729) | 0.694 (0.691, 0.699) | 0.705 (0.704, 0.706) |
| Fracture | **0.720 (0.719, 0.720)** | 0.673 (0.661, 0.682) | 0.411 (0.400, 0.421) | 0.289 (0.283, 0.298) |
| Lung Lesion | **0.886 (0.885, 0.886)** | 0.847 (0.844, 0.850) | 0.647 (0.640, 0.651) | 0.717 (0.711, 0.724) |
| Lung Opacity | **0.933 (0.933, 0.933)** | 0.895 (0.895, 0.896) | 0.777 (0.777, 0.778) | 0.794 (0.792, 0.796) |
| Pleural Effusion | **0.945 (0.945, 0.945)** | 0.935 (0.935, 0.936) | 0.809 (0.807, 0.811) | 0.762 (0.761, 0.765) |
| Pleural Other | 0.908 (0.908, 0.908) | **0.947 (0.945, 0.949)** | 0.915 (0.913, 0.917) | 0.652 (0.633, 0.683) |
| Pneumonia | **0.839 (0.838, 0.841)** | 0.752 (0.750, 0.756) | 0.606 (0.603, 0.614) | 0.705 (0.700, 0.709) |
| Pneumothorax | **0.894 (0.894, 0.894)** | 0.679 (0.678, 0.681) | 0.624 (0.621, 0.627) | 0.487 (0.485, 0.489) |
| Support Devices | **0.928 (0.928, 0.928)** | 0.852 (0.851, 0.852) | 0.796 (0.796, 0.796) | 0.753 (0.751, 0.756) |

**Table S6:** Additional selected concepts for building the CBM.

|  |  |
| --- | --- |
| • Large bibasilar atelectasis<br>• Heart size is chronically and severely enlarged<br>• Heart is moderately to severely enlarged though improved<br>• Two, now confluent left lower lobe and a consolidative right upper lobe opacity are all worsening<br>• Consolidation in the right midlung and left lower lung has worsened<br>• Left lower lobe consolidation more severe than right<br>• Interval worsening of bilateral interstitial pulmonary edema and vascular congestion<br>• Mild-to-moderate pulmonary edema bilaterally has worsened<br>• Widespread pulmonary edema has worsened<br>• Right middle and lower lobe atelectasis worsened<br>• Right lower lobe atelectasis has worsened substantially, perhaps lobar collapse<br>• Old lower left lateral rib fractures<br>• Compression deformity of the lower thoracic spine at least a year old<br>• Severe worsening of bilateral upper lobe predominant airspace opacities, right greater than left<br>• Bilateral opacities on the right and consolidation on the left have increased in extent and severity<br>• Small bilateral pleural effusions have worsened<br>• Bilateral pleural effusions improved<br>• Extensive fibrotic changes in the upper lobes bilaterally<br>• Chronic interstitial changes in the bilateral lungs<br>• Worsening multifocal pulmonary opacities, particularly within the right upper lobe and left lower lobe<br>• Right lower lobe consolidation which has worsened<br>• No right apical pneumothorax status post removal of right chest tube<br>• Improving small right apical pneumothorax<br>• Single-lead pacemaker placed with tip in left ventricle | • Heart size continues to be severely enlarged<br>• Heart size remains moderately to severely enlarged<br>• Heart chronically enlarged<br>• Worsening consolidation in the left lower lobe now involving the left upper lobe<br>• Severe bilateral perihilar pulmonary consolidation has worsened on the left, improved on the right<br>• Large consolidation in the left lower lobe has worsened<br>• Pulmonary vascular congestion has worsened to the point of early pulmonary edema<br>• Severe pulmonary edema worsened<br>• Bilateral atelectasis has slightly improved on the right and substantially worsened on the left<br>• Left lower lung atelectasis has worsened<br>• Bilateral lower lobe atelectasis has improved substantially<br>• Chronic right lateral rib fracture with adjacent atelectasis<br>• Severe widespread opacities have worsened in the lower lungs and left upper lobe but improved in right upper lobe<br>• Bilateral extensive asymmetric right greater than left lung opacities have worsened<br>• Bilateral lower lung consolidations have worsened<br>• Bilateral pleural effusion has worsened<br>• Most of the left upper lobe and the right lung apex chronically scarred<br>• Severe right lung scarring and concurrent emphysema<br>• Combination of emphysematous changes and scarring in bilateral lungs<br>• Right lower lobe and left upper lobe opacities have worsened<br>• Right apical pneumothorax reduced with chest tube in place<br>• Left apical pneumothorax resolved<br>• Transvenous right atrial biventricular pacer leads |

**Figure S1:**
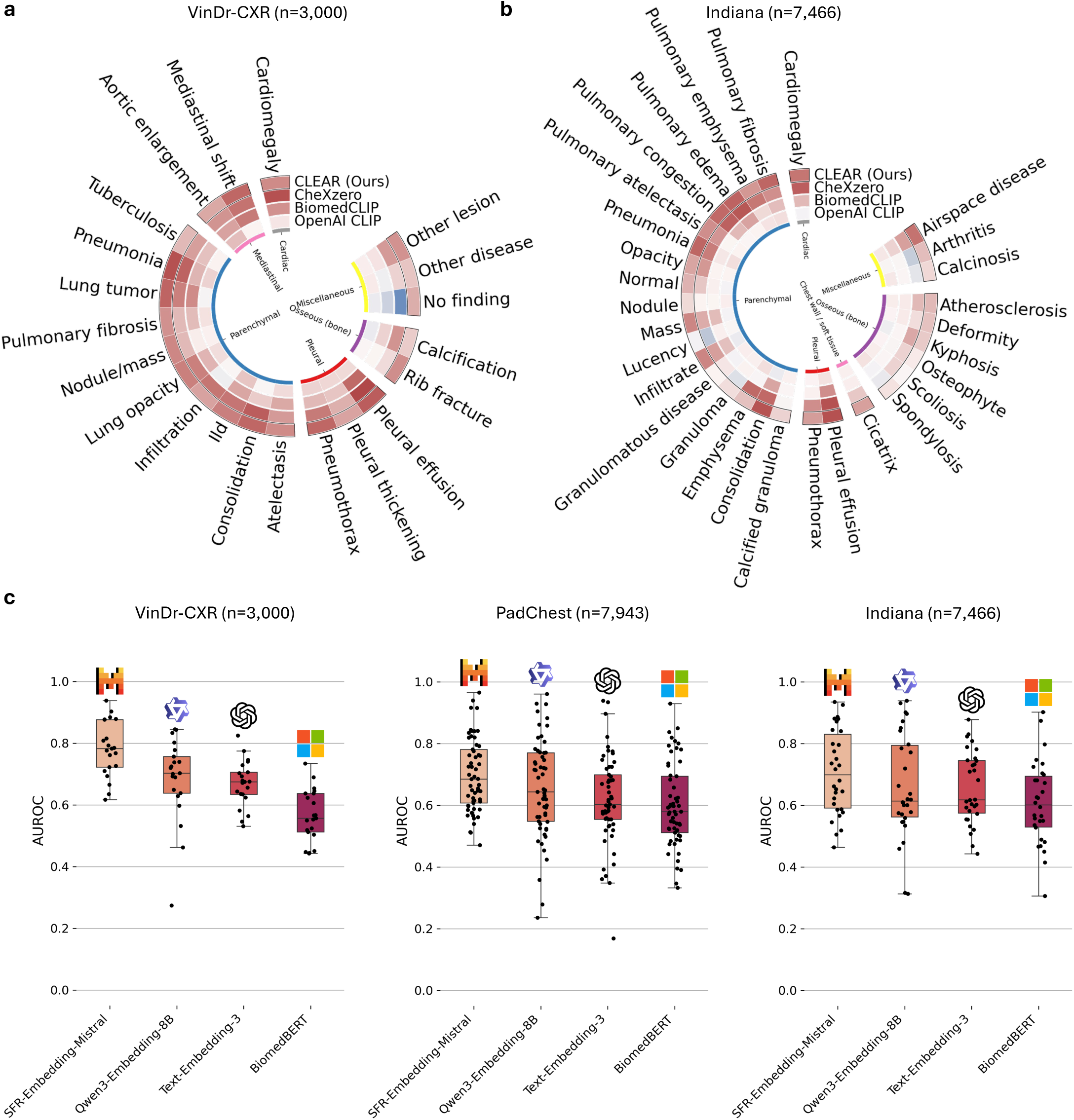
Comprehensive zero-shot performance and embedding model comparisons. **(a)**, Circular heatmap showing zero-shot AUROC performance on the VinDr-CXR dataset (n=3,000) across 21 radiological findings. **(b)**, Zero-shot performance on the Indiana dataset (n=7,466) across 30 radiological findings. **(c)**, Comparison of different LLM embedding models for CLEAR zero-shot classification. Box plots show AUROC distributions across all findings for CLEAR using different embedding backends: SFR-Embedding-Mistral, Qwen3-Embedding-8B, Text-Embedding-3-small, and BiomedBERT on the VinDr-CXR (left), PadChest (center, n=7,943), and Indiana (right) datasets. SFR-Embedding-Mistral consistently achieves the highest median performance. Boxes represent interquartile ranges with median lines; whiskers extend to 1.5× interquartile range; individual dots represent AUROC values for each finding.

**Figure S2:**
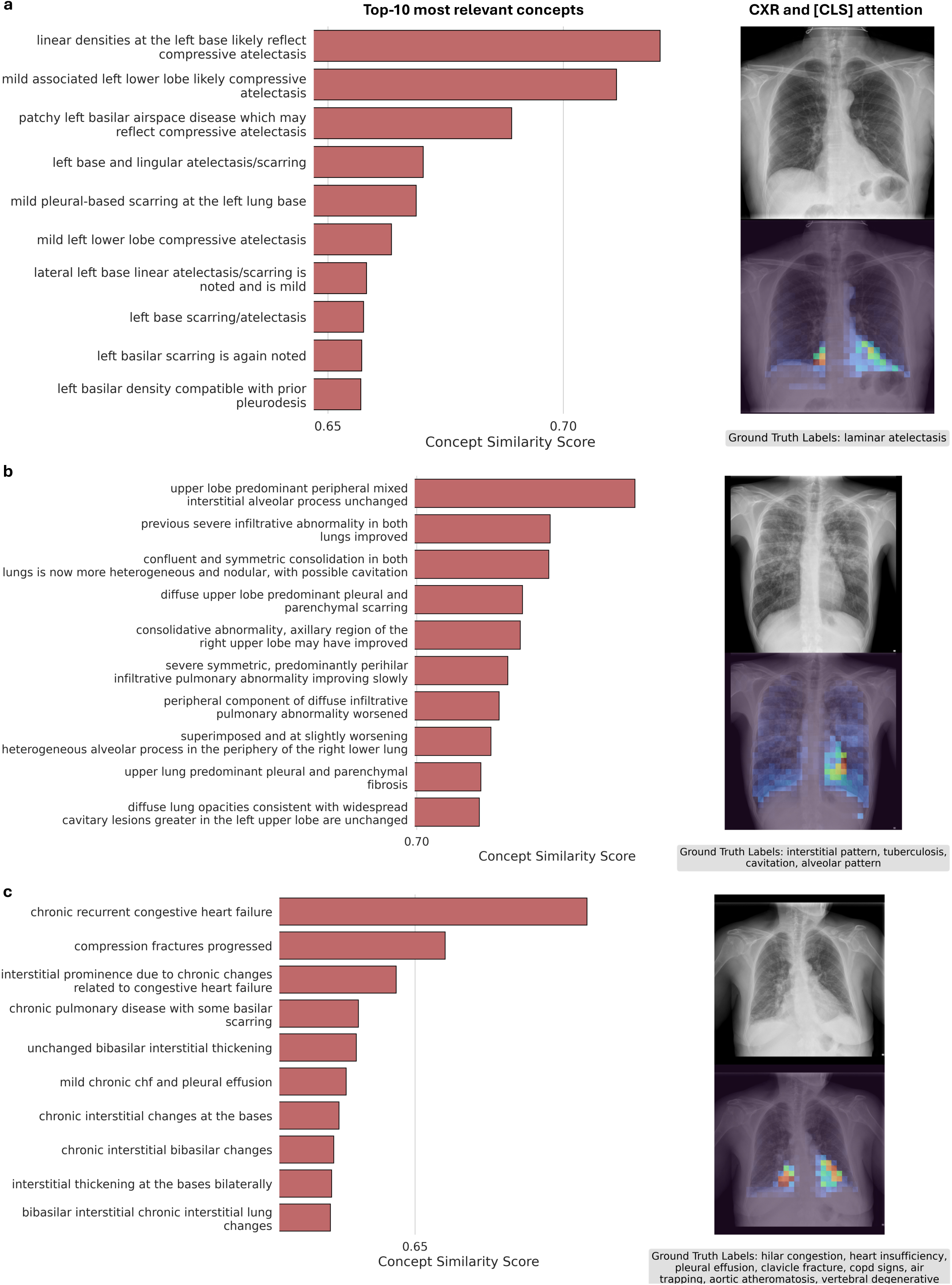
Concept auditing for zero-shot CXR classification. **(a)** Atelectasis detection showing top 10 relevant concepts with “linear densities at the left base likely reflect compressive atelectasis” scoring highest (0.70). The attention heatmap (bottom right) highlights the left lower lung region matching ground truth laminar atelectasis. **(b)** Multi-pathology case with CLEAR identifying interstitial-alveolar patterns. The attention heatmap focuses on bilateral upper lungs, consistent with ground truth: interstitial pattern, tuberculosis, cavitation, and alveolar pattern. **(c)** Congestive heart failure detection with “chronic recurrent congestive heart failure” as the top concept (0.65). The attention heatmap shows the bilateral distribution matching multiple cardiac-related ground truth findings.

**Figure S3:**
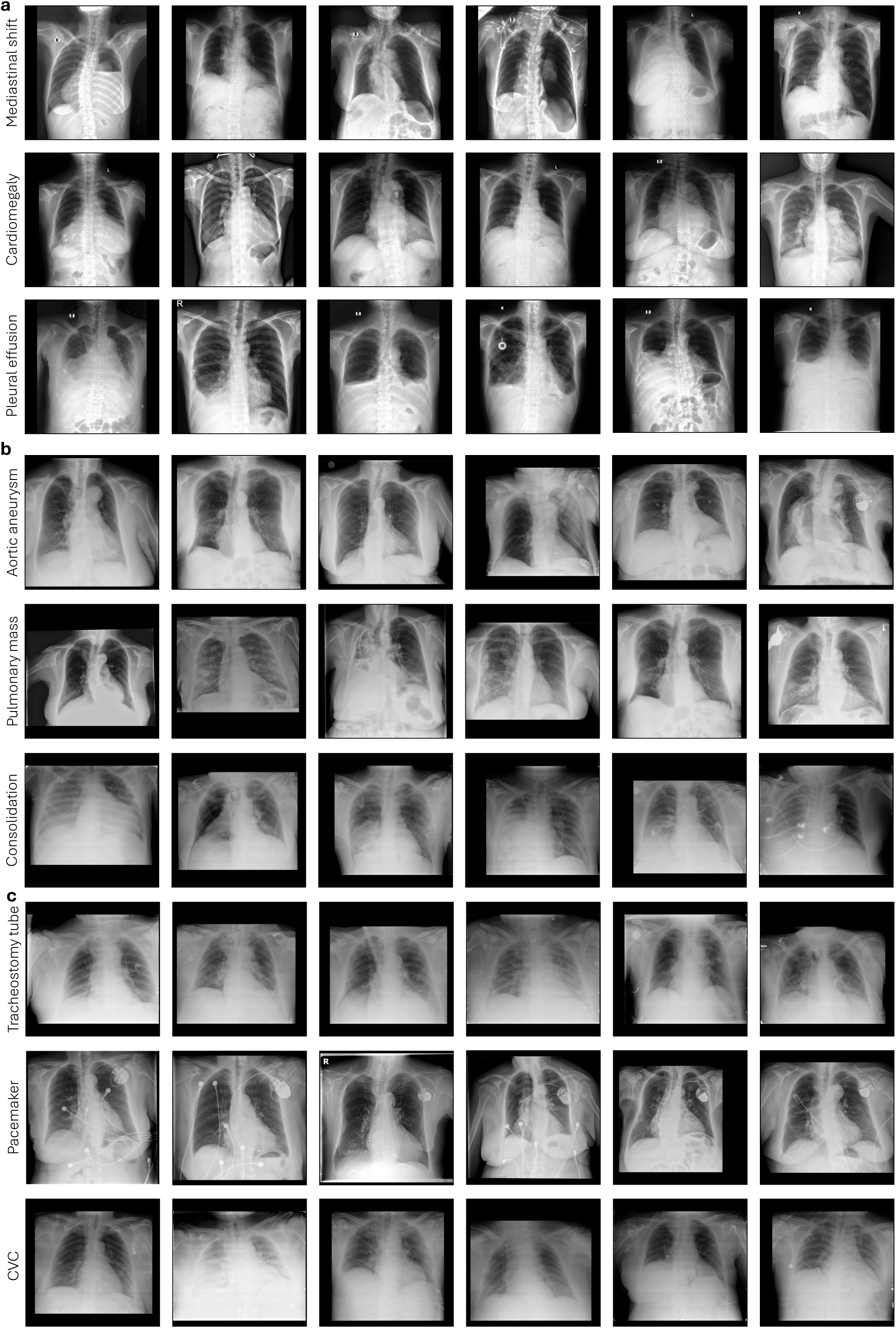
CXRs with high concept presence scores calculated using CLEAR. The concept presence score represents the degree to which a radiological observation is present in an image. Each row displays the six highest-scoring CXRs for selected concepts from the MIMIC-CXR dataset. **(a)** Anatomical findings: mediastinal shift, cardiomegaly, and pleural effusion. **(b)** Pathological findings: aortic aneurysm, pulmonary mass, and consolidation. **(c)** Medical devices: tracheostomy tube, pacemaker, and central venous catheter (CVC). Images demonstrate CLEAR’s ability to retrieve clinically relevant examples with visible radiological features corresponding to each concept.

**Figure S4:**
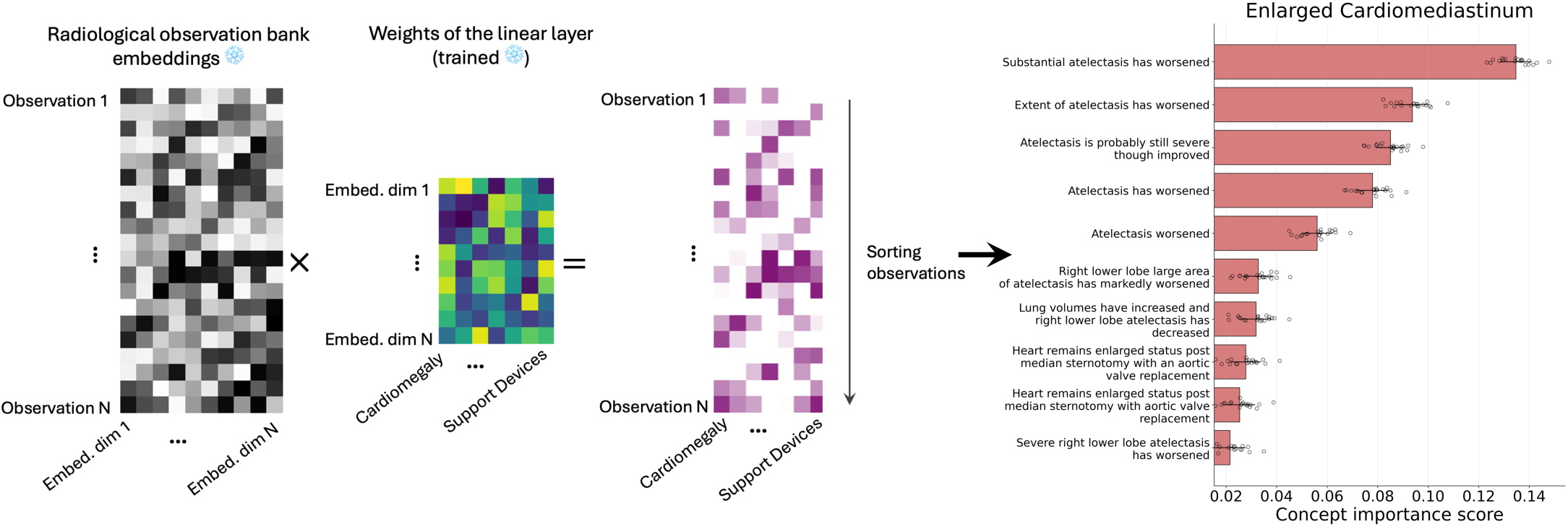
Concept importance analysis pipeline for supervised linear probing. The left panel shows the computational workflow: radiological observation bank embeddings are multiplied by trained linear layer weights to produce observation-specific importance scores for each diagnostic label. The resulting importance matrix is sorted to identify the most influential concepts. The right panel demonstrates the output for classification of enlarged cardiomediastinum.

**Figure S5:**
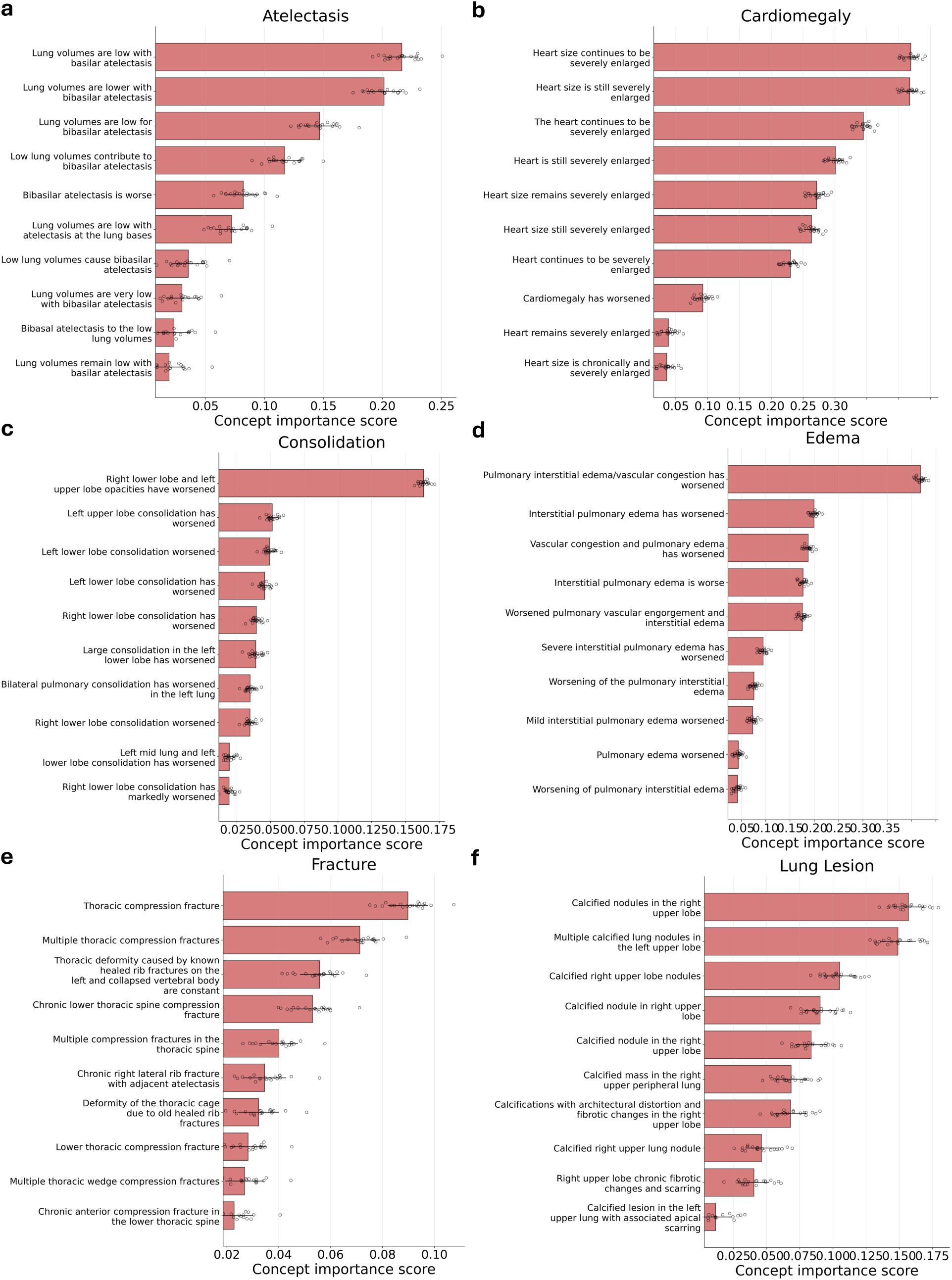
Concept importance output for atelectasis, cardiomegaly, consolidation, edema, fracture, and lung lesion classifications.

**Figure S6:**
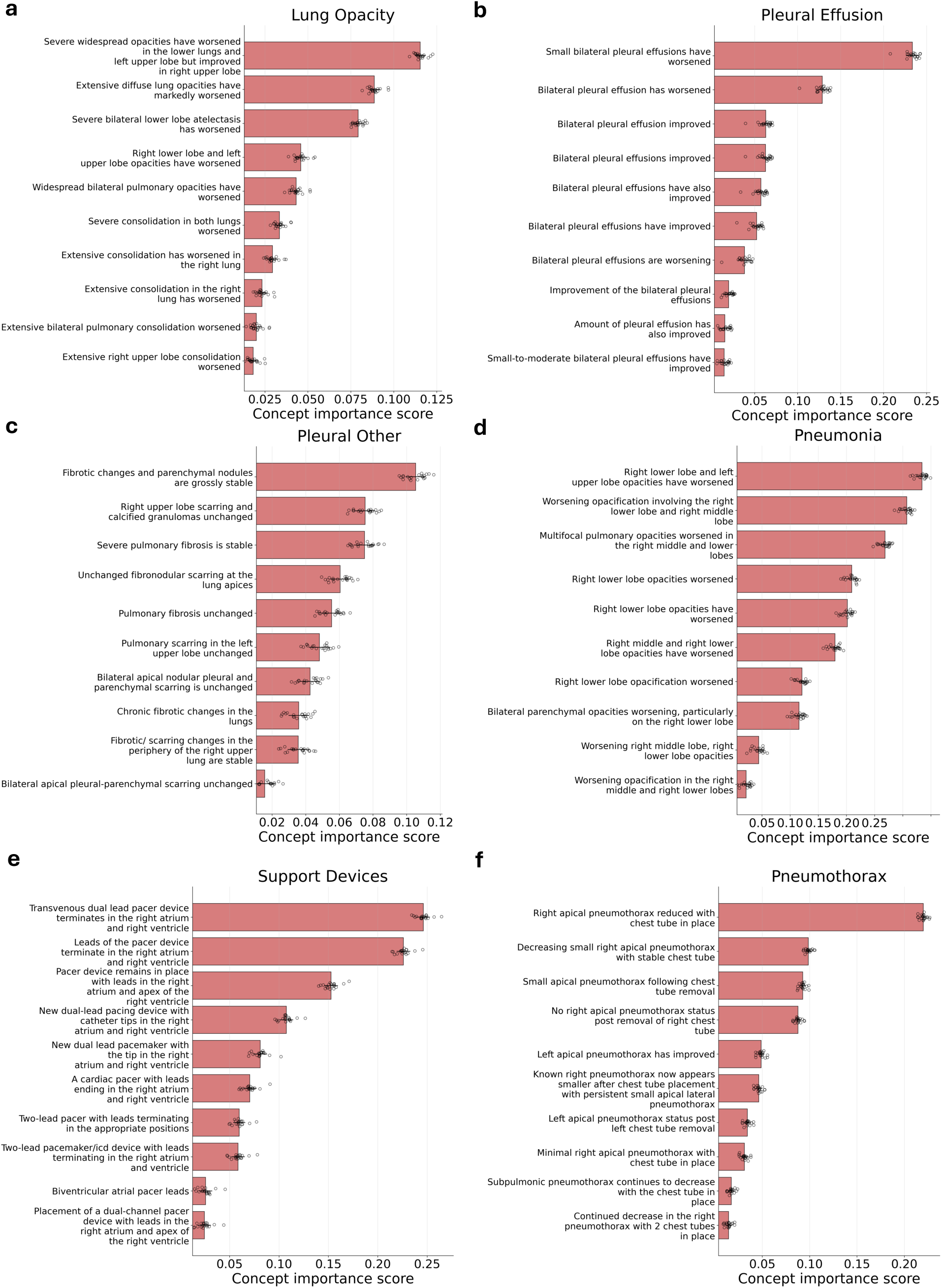
Concept importance output for lung opacity, pleural effusion, pleural other, pneumonia, support devices, and pneumothorax classifications.

**Figure S7:**
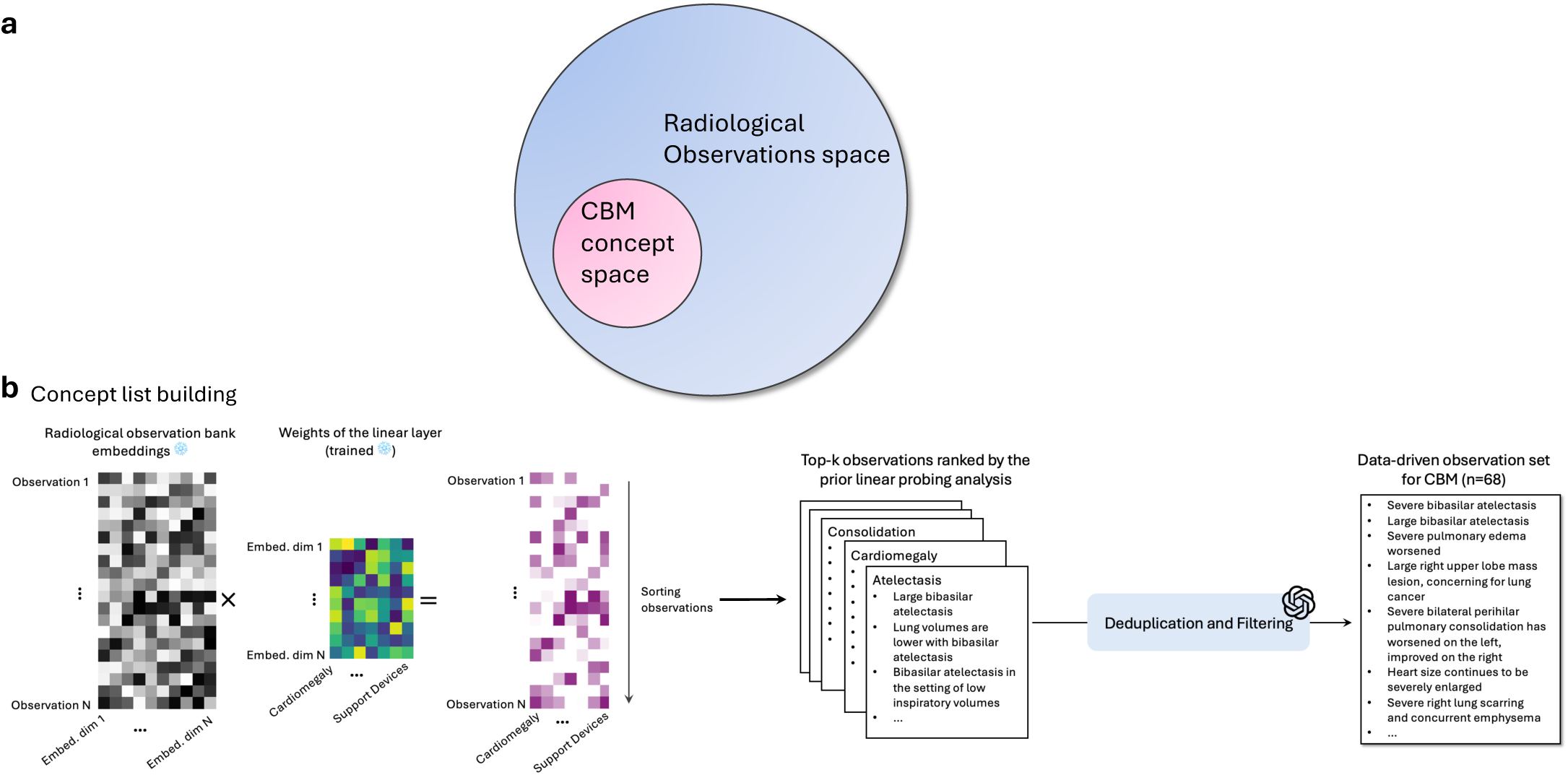
Data-driven concept selection for CBM construction. **(a)** Venn diagram illustrating that CBM concept space (n=68, pink) is a subset of the comprehensive radiological observations space (n=368,294, blue) extracted from MIMIC-CXR reports. **(b)** Concept list building pipeline. Left: Linear probing analysis identifies the most predictive observations for each diagnostic label by computing importance scores from trained weights. Center: Top-k observations ranked by importance across 13 pathologies (consolidation, cardiomegaly, atelectasis, etc.) are aggregated. Right: OpenAI GPT-4.1 performs deduplication and filtering to produce the final data-driven observation set of 68 high-quality concepts for CBM construction, including key findings like “Severe bibasilar atelectasis,” “Large bibasilar atelectasis,” and “Large right upper lobe mass lesion, concerning for lung cancer.”

**Figure S8:**
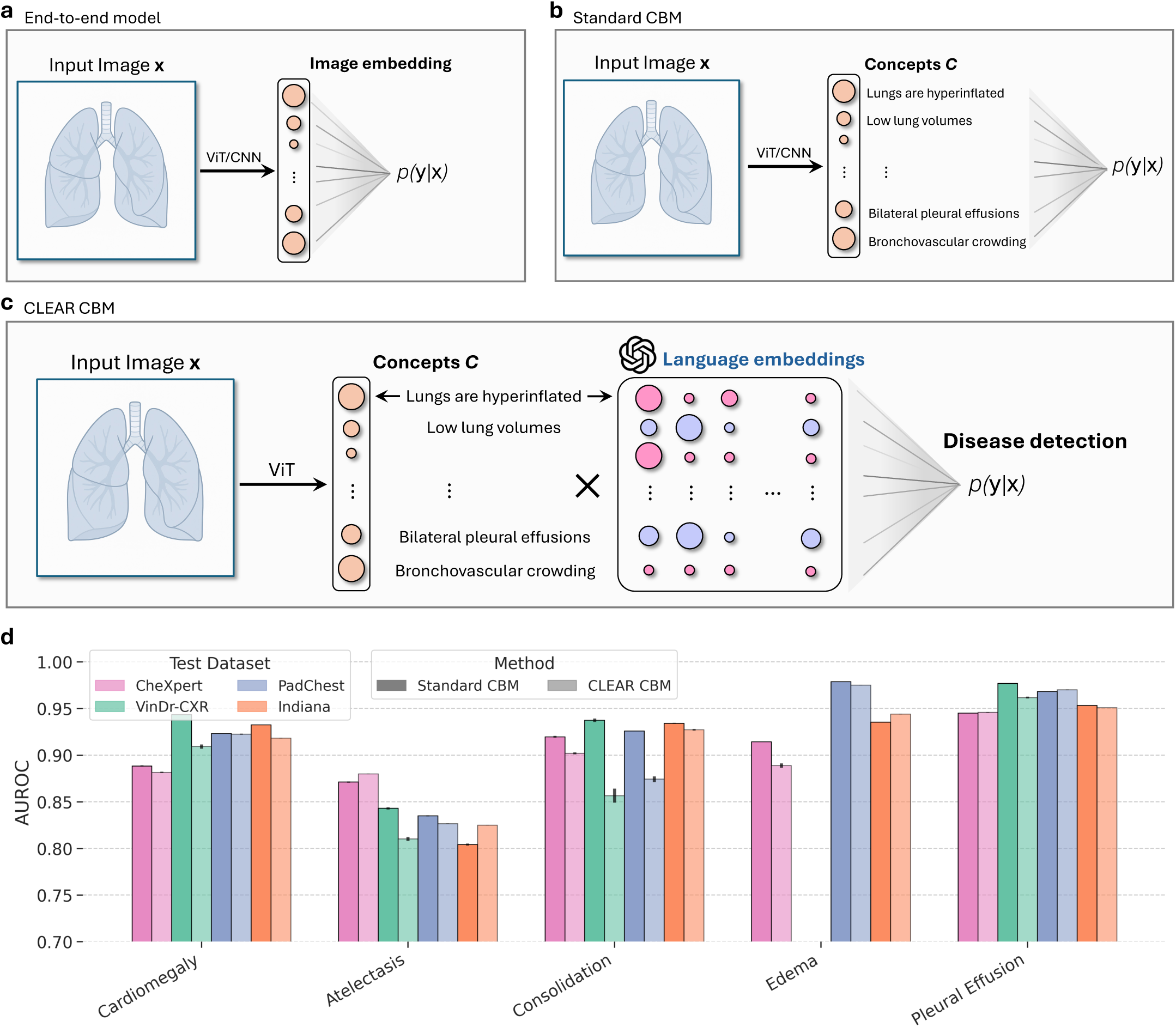
Comparison of model architectures and cross-dataset generalization. **(a)** The end-to-end model directly maps input CXR to disease predictions through opaque image embeddings using vision transformer (ViT) or CNN architectures. **(b)** The standard CBM enforces interpretability by first predicting concept scores (e.g., “Lungs are hyperinflated”, “Low lung volumes”) then mapping these to disease predictions through a linear classifier. **(c)** CLEAR CBM enhances the standard approach by incorporating LLM embeddings, multiplying concept scores with language embeddings to create semantically rich representations before disease classification. **(d)** Cross-dataset generalization performance. The bar charts show AUROC values for Standard CBM (dark gray) and CLEAR CBM (light gray) across four external test datasets (CheXpert, VinDr-CXR, PadChest, Indiana) for five CheXpert competition pathologies. Both CBMs achieve comparable performance, with AUROC exceeding 0.80 for cardiomegaly, atelectasis, consolidation, edema, and pleural effusion across most datasets, demonstrating robust generalization while maintaining interpretability.

